# Public participation in crisis policymaking. How 30,000 Dutch citizens advised their government on relaxing COVID-19 lockdown measures

**DOI:** 10.1101/2020.11.09.20228718

**Authors:** Niek Mouter, Jose Ignacio Hernandez, Anatol Valerian Itten

## Abstract

Following the outbreak of COVID-19, governments took unprecedented measures to curb the spread of the virus. Public participation in decisions regarding (the relaxation of) these measures has been notably absent, despite being recommended in the literature. Here, as one of the exceptions, we report the results of 30,000 citizens advising the government on eight different possibilities for relaxing lockdown measures in the Netherlands. By making use of the novel method Participatory Value Evaluation (PVE), participants were asked to recommend which out of the eight options they prefer to be relaxed. Participants received information regarding the societal impacts of each relaxation option, such as the impact of the option on the healthcare system. The results of the PVE informed policymakers about people’s preferences regarding (the impacts of) the relaxation options. For instance, we established that participants assign an equal value to a reduction of 100 deaths among citizens younger than 70 years and a reduction of 168 deaths among citizens older than 70 years. We show how these preferences can be used to rank options in terms of desirability. Citizens advised to relax lockdown measures, but not to the point at which the healthcare system becomes heavily overloaded. We found wide support for prioritising the re-opening of contact professions. Conversely, participants disfavoured options to relax restrictions for specific groups of citizens as they found it important that decisions lead to “unity” and not to “division”. 80% of the participants state that PVE is a good method to let citizens participate in government decision-making on relaxing lockdown measures. Participants felt that they could express a nuanced opinion, communicate arguments, and appreciated the opportunity to evaluate relaxation options in comparison to each other while being informed about the consequences of each option. This increased their awareness of the dilemmas the government faces.

## 1. Introduction

The Corona crisis is a vivid example of a critical juncture in the history of nations^1^. Following the outbreak of COVID-19, governments around the world took unprecedented measures to curb the spread of the virus, to protect high-risk groups and to prevent the overloading of health care systems. These government measures resulted in a range of unprecedented economic and social impacts^2^. Imposing such restrictions is a significant challenge for political leaders, who are pressured to decide under time constraints, often with limited knowledge of the future course of the crisis and the impacts of their decisions. While this is common to many types of disasters, pandemics are a rising tide, with prolonged uncertainty and accumulating cases. The potential mortality, morbidity, and life disruptions are difficult to predict, but waiting to act until the facts are certain is unacceptable to many political leaders^3^. From the beginning of the crisis up to the time of writing, one can observe a myriad of national and local responses to COVID-19, which differ in the composition of the policy mix but also in the timing and intensity of policy adoption^4^.

During periods of crisis and high uncertainty, the demand for scientific and technical expertise increases as governments and the public search for certainty in understanding problems and choosing responses^2,5^. In many countries, this creates a need for what is perceived as evidence-based policymaking, which signals to the public that decisions are being made based on reasoned and informed judgments that serve the public good, rather than special interests ^6^. Scientific and technical experts have become part of decision-making processes, as their names and images join political leaders as the face of how governments respond^2,5^. For instance, the Dutch prime minister Mark Rutte has said that he navigated this crisis guided by the knowledge of health experts from the Dutch Outbreak Management Team (OMT), members of which regularly participated in official press conferences. In Germany, the Chancellor received advice from two health experts: namely Christian Drosten, head of virology at Berlin’s Charité hospital and Lothar Wieler, the head of the government-funded Robert Koch-Institute^7^.

As scientific and technical experts become more prominent in defining problems and solutions during a crisis, the question of who is accountable for policymaking becomes more difficult to answer^2^. Moreover, the increased centrality of health experts in policy networks raises questions about the extent to which other types of expertise and interests (e.g. social and economic) are sufficiently heard and the extent to which the advice of health experts produces decisions that align with society’s preferences. In Germany, all virus-related policies made at the early stage of the pandemic were negotiated in an *ad hoc* way, largely bypassing the parliamentary system^7^. The core executives at the national and regional levels succeeded in rapidly concentrating decision-making power at the top of the pyramid. As Dostal^7^ concludes, the most important point of critique towards the German approach was the decision to limit the utilisation of expertise to a very small number of hand-picked experts. Avoiding ‘counter-expertise’ produced a form of tunnel vision among decision-makers, and many ostensibly ‘neutral’ expert recommendations involved value judgements and moral questions. Unsurprisingly, considerable differences in people’s attitudes towards COVID-19 policies are not only visible between countries but also within, especially across regions and age groups^8^.

When government decisions misalign with citizens’ preferences, society can correct political decisions by ‘voting with their feet’. For instance, the government of Serbia backtracked on its plans to enforce a second lockdown after major protests, and the Dutch government decided to close schools following protests, even though health experts from the Outbreak Management Team advised against school closure. However, democracy theorists would argue that such protests may not sufficiently represent the preferences of society at large^9-10^. In contrast to the examples in which citizens try to ensure that their voices are heard via protests, government-initiated public participation in COVID-19 policymaking has been notably absent^2,4,11-12^ despite being repeatedly recommended in health disaster response literature ^4,13-14^.

In a broad sense, the literature offers three rationales for involving citizens in crisis policymaking: the substantive, the normative and the instrumental rationale. The substantive rationale suggests that involving citizens will improve the quality of government decisions. Citizen participation allows a better evaluation of people’s preferences towards the impacts of government policies, which can provide input for governments to align their decisions with citizens’ preferences ^15-17^. Through a participatory process, the public may bring in new ideas, arguments, values and conditions that were not on the radar of (experts who inform the) decision-makers^18^. For instance, the celebrated concept of drive-through testing was a citizen’s idea^19^. The normative rationale asserts that involving citizens in policymaking is ‘the right thing to do’ in a democracy, as citizens should have a say in (governmental) decisions that will deeply affect their lives and society^20^. According to Lavazza and Farina^5^, health emergency policies that have strong ethical implications, deeply affecting people in very sensitive domains, should be participatory in character. Government-initiated participation in COVID-19 policies allows citizens to raise their voices in a more constructive and peaceful way than the protests in Serbia, Chile, Italy or the United States^21-22^. Finally, public participation exercises can be said to be motivated by an instrumental rationale when they aim to achieve a particular predefined end, such as increasing citizens’ acceptance of COVID-19 policies or restoring public trust. Greater public support for measures during a crisis can increase citizens’ compliance, which in turn is likely to increase the effectiveness of non-pharmaceutical measures^23-24^.

In the Netherlands, an attempt was made to involve 30,000 Dutch citizens in policy decisions regarding relaxing lockdown measures for the period of 20 May to 20 July, 2020 through a Participatory Value Evaluation (PVE). PVE is a preference elicitation method which can ameliorate the potential misalignment between government decisions and public preferences by measuring the latter in a large and diverse group of citizens. The essence of a PVE is that citizens can give advice on government decisions in an easy-to-access manner^25^; they are effectively put in the shoes of a policymaker. For example, in an online environment, they see: 1) which policy options the government is considering; 2) the concrete impacts of the options among which the government can choose and; 3) the constraint(s) that the government faces. Subsequently, citizens are asked to provide a recommendation to the government in terms of the policy options the government should choose, subject to the constraint(s). Individuals’ preferences over (the impacts of) policy options can be determined by feeding these choices into behaviourally-informed choice models^26^. The obtained preferences can be used to rank government policies in terms of their desirability.

The essence of a PVE can be illustrated with the following example. Suppose that a government considers four policy options (A, B, C and D). Each policy results in costs (let us assume 5, 10, 15 and 20 million euros) and a range of impacts (X, Y, Z). Suppose that the government faces a public budget constraint of 20 million euros. In this case, participants in the PVE will be asked how they would suggest the government allocate the 20 million euros over the policy options while being informed about the impacts of each of the policy options.

In this paper, we report the results of the PVE regarding the relaxation of lockdown measures in the Netherlands between 20 May to 20 July 2020. The primary goal of this paper is to show what type of insights a PVE can bring to policymakers and other stakeholders who have to decide on corona policies. A secondary objective of this paper is to improve understanding of the strengths and weaknesses of PVE in terms of involving citizens into crisis policymaking. To achieve this, we compare PVE with other methods and discuss the merits, in terms of the three rationales for public participation, of PVE in involving citizens in crisis policymaking. This comparison might provide policymakers with arguments as to why PVE is an appealing and feasible participatory method in times of a pandemic. That said, we do not aim to provide a conclusive answer to the question of whether PVE is better or worse than other participatory methods.

The remainder of this paper is organized as follows: section 2 discusses the three rationales for public involvement in crisis policymaking. Section 3 reasons why PVE is an attractive method for involving citizens in crisis policymaking by comparing the method with other participatory approaches. Section 4 discusses our methodology. Section 5 presents our results and section 6 provides a conclusion and discussion.

## 2. The rationale for active public involvement in crisis policymaking

Since the outbreak of COVID-19 in the first quarter of 2020, most governments have been operating in “emergency mode”. Scholars, pundits and journalists began warning at the beginning of the pandemic about risks like authoritarian power grabs, speeding up surveillance and other ‘temporary’ measures that will eventually outlast the pandemic^27-29^. Despite the fact that some political actors were indeed ready to exploit crises to change policies or institutions^30-31^, effective and agile, coordinated, consultative and collaborative approaches among government and non-government actors have taken the spotlight^24^. However, public participation in COVID-19 policymaking – using citizen advice in value-laden health policy decisions – has been notably absent^2,4,11-12^. Even routine forms of obtaining public input requiring minimal effort from public officials were hardly deployed. There have been a few instances of citizen involvement in COVID-19 policymaking in South Korea, Scotland, Belgium or Estonia, which we will discuss later in this section. However, even these examples only relate to the gathering of citizens’ ideas or evaluating attitudes towards new government measures. In the following passages, we present a range of prominent theoretical rationales for involving citizens in policymaking in general and crisis policymaking in particular. We classify the arguments according to Fiorino’s^32^ distinction between substantive, normative and instrumental justifications.

### 2.1 Substantive rationale

Due to the high urgency associated with decision-making during a pandemic, governments might easily overlook important details. For instance, some of the current policy plans might incorrectly assume that the public’s response will be guided by an almost exclusive focus on risk beliefs about the danger of the pandemic and the likelihood of being infected. Risks are evaluated within the context of people’s lives and priorities, and because of this, some risks may be judged as acceptable^33^. For example, low-income groups might have a stronger need to ignore self-quanrantine orders or travel restrictions in order to earn money to survive, since their relative earning losses are higher than for other income groups^33^. As studies have shown, the general public weighs pandemic policy decisions differently than professionals (who might have a tendency to view the world through a narrower lens)^13^. Hence, understanding how the risks and benefits of an intended policy are seen by the public will require input from groups outside the government and the health sector^14^. Through a participatory process, the public may bring in new ideas, arguments, values and conditions that were not on the radar of (the experts who inform) decision-makers^18^. In Scotland, such an exercise by its government led to over 4,000 ideas and 18,000 comments from citizens about the lockdown^34^. Citizens’ imaginations are not necessarily constrained by legalistic, bureaucratic or scientific views of disaster management, but have the potential to be a source of collective wisdom and capability to solve problems^14^. In South Korea, the government adopted some citizen-led strategies to fight COVID-19. For example, a student in that country developed a mobile application that citizens could use to access information on confirmed patients. Furthermore, as mentioned in the introduction, the concept of drive-through testing was also a citizen’s idea^19^. A potential caveat is that citizens’ input often needs to be produced in a short timeframe to have an impact on policy decisions. In crisis management, this window of opportunity can be rather small. Hence, once public officials have made up their minds, it can be too late for incorporating the publics’ input.

### 2.2 Normative rationale

When citizen participation is driven by a normative rationale, it is seen as ‘the right thing to do’. Citizens should have a say in governmental decisions when policies will affect their lives in significant ways^20^. Deliberative scholars argue that the far-reaching involvement of citizens in the design of public policies is especially important at the time of world-changing events like a pandemic. This is because elected officials have to take ethical decisions -ones that produce clear winners and losers which are beyond the mandate they received during elections held prior to the pandemic^12,35-37^. More importantly, the chances for greater victimization during a disaster or epidemic are unevenly distributed in society, as are the opportunities for enhanced safety. Economic means, social class, ethnicity and race, gender, and social connectedness are factors that often determine the extent of harm suffered^14^.For example, Hispanic Americans and African Americans have succumbed to COVID-19 in disproportionately higher numbers than the population as a whole^38^. Isolated individuals with few social ties are also more vulnerable to disasters ^39^. Including groups that might be un(der)represented in policymaking is therefore not only the ‘right thing’ to do, but such efforts also feed positively into the substantial rational of public participation; in many responses to COVID-19, policy effectiveness was reduced by ‘blindspots’ in otherwise well-performing systems due to failure to adequately care for vulnerable groups^31^. Moreover, the way we perceive the impact of government measures on the lives (and deaths) of others, will likely affect the way in which we sacrifice our personal freedoms for the benefit of the extended community. As studies and the protests in Serbia, Chile, Italy or the United States have shown, the general public weighs pandemic policy decisions differently than do professionals (who might tend to view the world from a narrower perspective)^13^.

### 2.3 Instrumental rationale

Public participation exercises can be said to be motivated by an instrumental rationale when they aim to achieve a particular predefined end, (e.g. increasing citizens’ compliance and trust). Greater public support for imposed lockdown measures can increase citizens’ compliance, which in turn is likely to increase the effectiveness of non-pharmaceutical measures^23-24^. Yet support for and compliance with a policy measure are difficult to model before that measure has been implemented^40^, since a myriad of individual, group, and subgroup responses to disease outbreaks affect attitudes and behaviour (e.g., perceived gender roles, generational differences, religious beliefs, partisanship, varying health literacy and education levels)^3,41^. Because of the high degree of uncertainty surrounding a new type of virus, people typically do not demonstrate the ability to fully process messages from the government. They must make quick judgments, based on emotion and a general feeling towards the government, in taking action^42^. This points to a circular relationship between how citizens evaluate their expectations towards their government and their evoked measures. In their survey before and after the lockdown in Western Europe, Bol et al.^43^ note that the expectation of policies was not enough to spur policy support; rather it is retrospective policy evaluation. It is worth emphasizing that,in some cases, the intrinsic sense of responsibility citizens feel might have a stronger explanatory power in terms of successfully suppressing COVID-19 outbreaks than do government measures. Unlike Taiwan or South Korea, Hong Kong’s success in fighting COVID-19 cannot be attributed to an executive that acted early, forcefully and with good governance backed by the people^44^. In an environment of low public trust and a lack of political legitimacy – which would together normally result in policy failure – Hong Kong’s citizens decided to organize their own COVID-19 response^45^.

Overall, involving the public in crisis policymaking is not something that government regularly do. Many policymakers remain sceptical about the contributions the public can make ^46-47^ and efficient policies^12^. Even in normal times, many public officials have come to view the public as something that should be kept at arm’s length rather than as a potential resource helping to produce better decisions on health policies^13^. However, if policies align with citizens’ preferences, then the likelihood of effective support from citizens will be greater ^4,12^. Hence, citizen ownership of exit strategies will be essential to ensure that solidarity prevails over discrimination^48^. And as the pandemic continues unabated, polls are showing waning public satisfaction with governments’ handling of the resulting crises^12^.

## 3. Positioning PVE against other participatory approaches

PVE can be conceived of as a participatory approach to effectively involve a large and diverse group of citizens in public policymaking^25^. At the same time, PVE is also a preference elicitation technique which can be used for the economic evaluation of government policy options^25-26^. Hence, PVE extends the substantive rationale for citizen participation by providing policymakers with insights into the economic costs and benefits of crisis policies. This section compares PVE with other participatory approaches to improve understanding of its strengths and weaknesses in terms of involving citizens in crisis policymaking. Note that we compare PVE with archetypes of other participatory approaches described in the literature and that we are aware of the fact that specific versions of an approach might exist with a different set of strengths and weaknesses. Moreover, we focus here on public participation in crisis policymaking, not in the overall management of a public health crisis, which can also include other forms of participation. The literature provides a range of criteria for defining whether a method or a process can be conceived as a ‘participatory approach’, and sometimes these criteria can be quite restrictive^49^. In the present paper, we classify a method as a participatory approach when it is explicitly used as a public consultation preceding a governmental policy decision.

### 3.1 Mini-publics

The literature offers a range of participatory methods to involve citizens in the design and evaluation of public policies which centre around deliberative mini-publics; examples include citizen assemblies and consensus conferences. In essence, a mini-public is a demographically representative sample of the population, small enough to genuinely deliberate, and representative enough to be genuinely democratic^50^. A mini-public generally consists of around 15 to 100 randomly selected citizens (there are examples with 500) who, enabled by an independent facilitator, collectively provide advice on a policy issue^10^. Citizen assemblies are one example of a mini-public that has been successful in dealing with divisive and highly politicised issues such as same-sex marriage, abortion and decarbonisation measures. The purpose of a citizen assembly is to employ a cross-section of the public to study the options available to the government on certain questions and to propose answers to these questions through dialogue and the use of various methods of inquiry such as directly questioning experts^51^.

The basic reasoning behind deliberative approaches is that a diverse and inclusive group of citizens, if given adequate information, resources and time to deliberate on a given topic, can produce an informed judgement. The Deliberative Democracy Consortium defines deliberation as “an approach to decision-making in which citizens consider relevant facts from multiple points of view, converse with one another to think critically about options before them and enlarge their perspectives, opinions, and understandings”^52^. Participants must consider a question from multiple viewpoints, exchange perspectives, opinions, and understandings and think critically about all possible options. The emphasis is to engage participants from the affected population, without excluding social groups or marginalised views^10^.

The main downside of deploying deliberative approaches for involving citizens during a pandemic is that such processes generally take a lot of time. The biggest logistical task remains the selection process, which must deliver a representative sample of a given population, as well as a range of experts from different disciplines, with different perspectives on the matter in question^12^. Moreover, participants must take time to educate themselves and exchange viewpoints. This is tricky because policy questions during a pandemic are highly volatile, and governments have to respond quickly to new developments. For instance, the Irish Citizen Assembly on Abortion took more than a year to produce final recommendations and the French citizen convention on climate issues lasted for six months. And even though the actual face-to-face deliberations of the Public Engagement Project on Control Measures for Pandemic Influenza in the United States^53^ lasted one month, the project’s duration from planning to final report lasted eight months. Another issue with deliberations is that they are more effective offline, with participants able to engage in face-to-face interactions. This is relatively difficult in times of social distancing measures that were especially stringent at the peak of the pandemic. Furthermore, deliberation is usually carried out in small groups to ensure high-quality discussions, since this is unlikely to be possible with large groups^50^. This restricts the extent to which the public may bring in new ideas, arguments, values and conditions that were not on the radar of experts and decision-makers. Indeed, in a public health crisis, the aim should be to gather and circulate as many views as possible, to ensure that policymakers are as familiar as they can be with the social landscape that any resultant policy will need to be built upon^12,54^.

Furthermore, as Goodin^55^ argues, mini-publics should be deployed only if the views they reach are representative or at least an accurate reflection of those that would have been reached by a larger group had similar processes been feasible at that scale. It can therefore be argued that a group of 100 citizens might be too small to be able to provide a representative picture of the population’s preferences regarding a pandemic which is responsible for unprecedented and multi-dimensional impacts.

Finally, due to the participation of small groups, the number of citizens who will have increased their awareness through participation is also relatively limited. The way citizens perceive the impact of government measures on the lives (and deaths) of others will be mostly limited to the participants. During the deliberation on the US pandemic influenza policy in 2007, the exercise may itself have served as a trust-building exercise for the 260 citizens and the 50 government officials and stakeholders who participated. However, it was concluded that greater use of this method may be needed to assure both groups of the soundness of plans during an influenza pandemic^53^.

### 3.2 Referendum

An alternative approach for involving citizens in the evaluation of public policies is the referendum. The referendum reaches a larger and more diverse group of citizens because of its low ‘barrier to entry for participating’. The only effort that citizens have to expend is in casting their vote, Moreover, organizing referenda can be an opportunity to restore the legitimacy of public decision-making^56^. The lockdown measures imposed by governments were not discussed during previous election campaigns. Thus, citizens were not given the opportunity to take them into account when transferring authority to their elected representatives, something for which a referendum can correct. However, the referendum has several disadvantages in its application to crisis policymaking. Firstly, organising a ballot during a pandemic demands a great deal of time and effort in preparation. Secondly, citizens are only asked to vote ‘for’ or ‘against’ a proposal in a referendum, which prevents the public from expressing the kind of nuanced opinions which can enhance policy proposals or modify them to vulnerable groups. This is even more problematic if it neglects to address the subsequent policy implications of the choices on offer (for example, if the UK votes to leave the EU, how should it go about doing so?). Multi-dimensional policy issues such as those that arise during a pandemic generally do not lend themselves to a simple ‘yes’ or ‘no’ response. As Offe^57^ puts it, holding referenda on substantial yet unknown long-term results will only encourage the accountability-free expression of poorly considered mass preferences and de-emphasize requirements of consistency, compromise-building, and the reflection on consequences. Moreover, a referendum does not allow citizens to transmit new ideas, arguments, values or conditions to decision-makers. Finally, if the outcome of a referendum is considered to be binding, this would limit a government in responding quickly to new scientific insights or to new developments during a highly volatile pandemic. Therefore, depending on the qualification requirements and on the kinds of policy proposals that are open for the ballot, referenda are mostly used to guide long-term strategic government decisions, rather than short-term measures and regulations^58^.

### 3.3 Opinion poll/survey

Governments also consult citizens through opinion polls, in which they ask them about the extent to which they support a certain policy or to rate several policy options. Such methods can be deployed rapidly and often make use of large randomised and representative panels, or are open for anyone to participate, such as ‘the big Corona study’^59^ of the Universities of Antwerp, Hasselt and KU Leuven. However, similarly to the referendum, the questions that are asked in these opinion polls are frequently too generic to be of much policy relevance. Questions such as “do you support the lockdown” or “where should wearing face masks be obligatory” may provide policymakers with a quick understanding of public opinion regarding these topics. However, polls do not provide a deeper insight into the extent to which people value one potential policy over another and how their preferences for a certain policy option are influenced by its (societal) effects^60^. Nor do such questions provide an opportunity for participants to experience the dilemma of the policymaker during a pandemic. Hence, the ability of public polling to inform policymakers is generally limited, especially when the impacts of policy trade-offs on citizens’ lives are not made visible.

### 3.4 Participatory Budgeting

A relatively new member of the family of direct democracy institutions is participatory budgeting (PB)^61-62^. The essence of PB is that non-elected citizens are involved in the allocation of designated parts of the public budget^63^; they do this by selecting a portfolio among the many portfolios that are possible within the budget. PB processes generally attract large and diverse groups of citizens because the barriers to entry are low. Putting large groups of people in the shoes of a policymaker might raise their awareness of intricate government dilemmas and may help set realistic expectations about the impacts of public health measures. It can be argued that PB constitutes a balancing point between the high barriers to entry and running time of mini-publics and the overly simplistic referendum/opinion poll. However, the subject of the exercise of a PB is pretty clear: to divide up a public budget. In contrast, during a pandemic, money is far from the only relevant scarce public resource over whose use a government needs to establish priorities.

### 3.5 Participatory Value Evaluation

Participatory Value Evaluation (PVE) closely resembles PB in the sense that citizens’ optimal policy portfolios are elicited given a constraint faced by the government in allocating public resources. A fundamental difference between the two methods is that the design of a PVE can adopt other constraints than only public budget (e.g. sustainability targets, maximum pressure on the health care system). PVE has three practical advantages over PB in the sense that in theory these characteristics can also be incorporated in a PB. First, a PVE explicitly communicates to participants that they can advise against allocating public resources to the proposed policy options. That is, participants are asked whether they advise the government to allocate any resources at all, and if so, which policy options they would recommend. Hanley et al.^64^ assert that such an experimental design, in which the baseline is clearly presented, will yield accurate estimates of the impacts of the implementation of policy options on citizens’ welfare. A second practical advantage is that insights can be obtained from a PVE regarding the extent to which preferences for policy options are affected by impacts of policy options by using sensitivity analyses (we will provide examples in section 5.3). That is, analysts can identify how the desirability of policy options is affected by changes in impacts. Third, in a PVE, the written motivations that participants use to explain their choices provide policymakers with insights in people’s arguments, concerns and values.

A difference between PVEs and mini-publics is that PVE experiments are based on individual preference formation. That is, respondents are provided with information on the policy alternatives they are meant to choose from, but they study this information individually, without the opportunity to ask questions or discuss. This approach has been criticised for implicitly or explicitly assuming that people have pre-formed preferences for quite abstract issues, such as COVID-19 lockdown measures, even when they do not have any relevant real-life experience^65^, or they are assumed to be able to form preferences in private based on informational material provided within the survey^66^. Various scholars argue that discussions with others and the opportunity to ask questions are decisive for preference formation, as preference formation is an inherently social and dynamic process^66-67^.

Table 1 provides a comparison between PVE and other participatory approaches on four dimensions. The goal of this comparison is to provide arguments as to why PVE could be an appealing and feasible participatory method in times of a pandemic. The purpose is not to provide a conclusive answer to the question of whether PVE is better or worse than other participatory methods.

**Table 1:** comparing PVE and other participatory approaches

|  | **Practical feasibility during pandemic** | **Substantive rationale for participation** | **Normative rationale for participation** | **Instrumental rationale for participation** |
| --- | --- | --- | --- | --- |
| **Mini-public** | -Setting up mini-publics takes a lot of time, which is inconvenient during a pandemic as policy questions are highly volatile and governments have to respond quickly to new developments.<br>-Mini-publics are more effective offline, which is complicated in times of social distancing. | +Deliberation in mini-publics positively affects the quality of information, preferences and arguments, and brings about new ideas that have not been on the radar of decision-makers.<br>-Mini-publics only work well with a group of around 100 citizens, which might be too small to be able to provide a representative picture of the population's preferences regarding a pandemic which will have unprecedented, uncertain and multi-dimensional impacts. | +Mini-publics perfectly align with the ideal of deliberative democracy. A proposed policy-change of a mini-public can be seen as more legitimate if citizens feel represented by the selected members. | -Members of mini-publics are often aware of the impacts of the policy decisions which they are advising on. The number of citizens who increase their awareness through participation is, however, relatively limited for those involved in a mini-public. |
| **Referendum** | -Organising a ballot during a pandemic requires a lot of preparatory time and is a costly endeavour. | -Referenda do not provide information about the extent to which citizens value (the impacts of) policy options.<br>-Referenda do not allow citizens to transmit new ideas, arguments, values or conditions to decision-makers.<br>-If the outcome of a referendum is considered binding, this would limit a government in responding quickly to new scientific findings or to new developments during a highly volatile pandemic. | +The referendum reaches a large and diverse group of citizens because of its low 'barrier to entry for participating'.<br>+Organizing referenda can be an opportunity to restore legitimacy in public decision-making<br>-Citizens are only asked to vote 'for' or 'against' a proposal in a referendum and it therefore does not allow the public to express nuanced opinions<br>on multi-dimensional policy issues. | +Depending on the clarity of the potential impacts as well as on the deliberative quality of the public debate preceding the ballot, a referendum can raise awareness on a policy issue for large groups of citizens. |
| **Opinion poll/survey** | +Can be deployed rapidly<br>+Online environment unimpacted by social distancing | +Provide policymakers with a quick understanding of public opinion regarding these topics.<br>-Do not provide a deeper understanding of the extent to which people value a potential policy over another and how people's preferences for a certain policy option are influenced by its (societal) effects. | -Do not provide an opportunity for participants to experience the challenges faced by policymakers during a pandemic. | -Polling's ability to raise awareness is generally limited. |
| **Participatory Budgeting** | -Focuses on the allocation of a public budget, which in a pandemic is not the only relevant scarce public resource whose use a government needs to prioritize.<br>+Can be deployed rapidly | +Provides insights into the allocation of a constrained public budget towards policy option(s).<br>-Does not allow citizens to transmit arguments, values or conditions to decision-makers. | +The literature portrays PB as an innovative operationalization of direct democracy. | +Putting large groups of people in the shoes of a policymaker might raise their awareness of intricate government dilemmas and may help set realistic expectations about the impacts of healthcare measures. |
| **Participatory Value Evaluation** | +Can be deployed rapidly<br>+Online environment unimpacted by social distancing | +Provides insights about the allocation of constrained public resources towards (the impacts of) a predetermined set of policy option(s)<br>+Outcomes can be used for the economic evaluation of policy options.<br>+Allows citizens to transmit new ideas, arguments, values and conditions to decision-makers.<br>-Quality of preferences that people express is probably lower than those expressed after deliberation (such as is the case in mini-publics). | +Provides an opportunity for participants to advise their government after experiencing a dilemma faced by policymakers.<br>+Allows citizens to express preferences about the distribution of benefits and burdens that accrue from government policies. | +Putting large groups of people in the shoes of a policymaker might raise their awareness of intricate government dilemmas and may help set realistic expectations about the impacts of health care measures. |

In conclusion, there are various reasons why PVE could be an appealing participatory approach for involving citizens in policy decisions during a pandemic. In terms of its practical feasibility, citizens can participate in a PVE online, which is appealing in times of social distancing. Moreover, a PVE can be deployed rapidly, which is important during a pandemic as governments have to respond quickly to new developments. The design of a PVE can also adopt other constraints than just the public budget, which is a key benefit compared to PB. In terms of improving the quality of decision-making (substantive rationale for participation), PVE provides information to policymakers about the extent to which the desirability of policy options is affected by the impacts of those options. It also allows citizens to transmit new ideas, arguments, values and conditions to decision-makers. From a normative point of view, a benefit of PVE is that it enables citizens to participate in multi-dimensional policy issues that do not lend themselves to a simple ‘yes’ or ‘no’ or the allocation of a constrained amount of public budget. From an instrumental point of view, letting citizens experience intricate government dilemmas improves their understanding of the social, health and economic impacts of proposed measures, which might also subsequently increase levels of acceptance and compliance.

## 4. Methodology

Before presenting the specifics of the PVE, section 4.1 compares PVE with contingent valuation (CV) and discrete choice experiments (DCE), which are two related preference elicitation techniques that can be used for the economic evaluation of government policy options. In this section, we also provide arguments as to why we selected PVE instead of these two other elicitation techniques for studying Dutch citizens’ preferences over the relaxation of lockdown measures. In section 4.2, we discuss the choices that we made in the design of the PVE. In section 4.3, we discuss the analysis techniques that were used in this study.

### 4.1 Comparing PVE with CV and DCE

CV is a valuation method based in surveys, designed to create a hypothetical market for public goods, and determine the amount of money that people would be willing to pay (willingness-to-pay, WTP) or accept as compensation (willingness-to-accept, WTA) for specific changes in the quantity or quality of such goods^68^. CV is a popular method in the field of environmental economics for answering questions such as how to value changes in environmental quality^69-70^. In the CV survey, participants first receive a detailed description of a proposed government project as well as the consequences of the project. Then, they are asked whether they are willing to pay a predetermined amount of money, commonly presented as a one-time tax, to finance the implementation of the project. The CV survey is completed by a representative sample of the population, while varying the amount of money required to implement the project. In this way, it is possible to obtain an estimate of the mean WTP of the population through econometric techniques^71^. In turn, this mean WTP estimate represents a measure of the welfare change generated by implementing the government project^72^.

While CV seems to be an effective method for determining the value of a whole project, its applicability as a preference elicitation technique is limited. Crucially, it is not possible to determine the extent to which different characteristics of the project (hereafter “attributes”) affect these preferences. Hence, CV is an attractive preference elicitation technique if the government wants to know society’s aggregate willingness to pay for one specific relaxation option, but from a CV it is not possible to infer how the aggregate willingness to pay for a particular relaxation option is affected by its impact on COVID-19 related deaths, physical injuries and mental injuries respectively.

An alternative for CV is to use a discrete choice experiment (DCE). The core idea behind DCEs is that individuals’ preferences for a government project are established by decomposing the project into separate attributes and different specifications of these attributes (referred to as ‘attribute levels’)^73^. The relative importance of these attributes can be empirically assessed by presenting respondents a series of choice tasks in which they are asked to choose a preferred alternative (in this case a specific relaxation option for lockdown measures) from a set of two or more alternatives with varying combinations of attribute levels^74^. By collecting the choices of a large group of respondents, statistical methods known as discrete choice models^75^ are used to estimate the preferences of individuals for policy options and attributes. These models have a solid foundation in random utility theory^76^, allowing researchers to compute welfare measures for changes in the quantity or quality of the attributes, and to determine the WTP of individuals for these changes^71^.

The literature distinguishes between labelled DCEs and unlabeled DCEs^74^. Unlabeled DCEs only focus on estimating people’s preferences for the concrete attributes of policy options and do not specify policies in terms of their nature (e.g. re-opening the hospitality industry or relaxing restrictions for young citizens), whereas labelled DCEs also specify the policy options which are evaluated by respondents in terms of their nature. The advantage of unlabeled DCEs is that it allows policymakers to use outcomes for the assessment of (combinations of policies), including those that are currently not on the table but might be considered in later phases of the crisis. A recent application of an unlabelled DCE to study the preferences for the relaxation of COVID-19 measures is provided by Chorus et al.^60^. An advantage of labelled DCEs is that it allows participating citizens to express their preferences towards a particular relaxation option regardless of the impacts that are included in the DCE.

Labelled DCE and PVE are closely related in the sense that both preference elicitation techniques allow individuals to express preferences towards specific policies as well as policy impacts. A first fundamental distinction is that participants in a DCE express preferences through selecting *a single policy option*, whilst participants in a PVE can select a *bundle of policy options*. Hence, participants in a PVE can evaluate policy options in relation to each other. Participants in a PVE can select one policy option or none of the options (just as in a DCE with an opt-out option), but – unlike in a DCE – they can also choose two or more options. A second fundamental distinction is that participants in a PVE express preferences not only towards specific government policies, but also towards the allocation of scarce public resources. Participants make a continuous choice regarding the extent to which they think that public resources should be allocated and discrete choices as to whether or not to include specific policy options in the bundle that they recommend to the policymaker. Participants in DCEs generally do not receive information concerning the scarcity of public resources and when such information is provided, participants are asked to recommend a single policy option from a set of policy options that all require the same investment of public resources^77-78^.

Whether or not a policymaker should choose PVE, (labelled or unlabelled) DCE or CV as a preference elicitation technique depends, in our view, on the policy question that should be answered. CV is an appealing technique when a policymaker wants to know whether a single relaxation option should be implemented; an unlabelled DCE is an appealing technique if the policymaker wants to know how individuals value the impacts of known and unknown relaxation options; labelled DCE is a promising elicitation technique when a policymaker wants to obtain information concerning people’s preferences towards both the impacts of policy options as well as the options in and of themselves; finally, a PVE is appealing when policymakers want to know people’s preferences regarding the extent to which scarce public resources should be allocated towards the (impacts of) a predefined set of options.

After the first wave of the pandemic had reasonably flattened, leaders in the Netherlands began contemplating about lifting lockdown policies. In the first week of April 2020, the research team heard from Dutch policymakers that they were expecting a major decision to be made in May. This decision concerned the ways in which the lockdown measures could be relaxed without overloading the healthcare system. Policymakers told the research team that they were considering various relaxation options which would have a range of societal impacts. We found PVE to be the most suitable preference elicitation method for this decision problem, as it concerned the allocation of scarce public resources (available capacity of the health care system) towards (the impacts of) a predetermined set of policy option(s).

### 4.2 Design of the PVE

We started on 9 April, 2020 with the design stage of the PVE via an online brainstorm with policymakers and researchers from the RIVM (the Dutch National Institute for Public Health and Environment), the Ministry of Health, Welfare and Sport and the Ministry of Finance about the relaxation options and impacts that they were considering. Based on this brainstorm, we compiled a shortlist of relaxation options and their impacts, which we discussed with various academics. In these meetings, we inquired as to whether we had overlooked important relaxation options and whether they could help us with providing information regarding the order of magnitude of the impacts of these strategies. For instance, we spoke with several epidemiologists to learn about the effect of relaxation options on the available capacity of the healthcare system as well as the number of deaths and people with permanent injuries caused by COVID-19. Moreover, as a result of these meetings, we included the option “All restrictions lifted in the Northern provinces”, as some academics we spoke with found this an attractive option and argued in the public debate for its inclusion^79^. These researchers considered this a promising approach, since at the time that the PVE was conducted there were only a few infections in these provinces; this made it easier to keep infection levels low through testing and tracing. In addition, we decided to split the attribute ‘increase in the number of deaths caused by the relaxation option’ into ‘additional deaths of people of +70 years’ and ‘additional deaths of people younger than 70 years’ as various academics we consulted found it interesting to know whether Dutch citizens weigh the increase of mortality risk differently between these two age groups.

Based on the information and feedback we received from policy makers and academics, we selected eight relaxation options and sent a draft version of the PVE to the policymakers for feedback. In the meantime, the research team collected reports and media content to describe the eight relaxation options in the PVE and to provide estimates of the attribute levels. For instance, we used projections regarding the increase in the number of people with lasting physical injuries caused by postponed operations^80^, data on the increase in domestic violence resulting from the corona crisis in the United Kingdom^81^, information on domestic violence in the Netherlands prior to the crisis^82^ and estimates concerning bankruptcies, unemployment and income loss^83-85^. We integrated this information and the feedback of policymakers into a new draft version of the PVE and this experiment was tested by a convenience sample of 80 respondents. We incorporated this feedback into the final version of the PVE.

In the PVE, participants were invited to advise the government on which lockdown measures should be relaxed between 20 May and 20 July 2020. They were asked if the government should relax lockdown measures during this period at all and, if so, which relaxation option(s) should be favoured. In an online environment (here the [original Dutch version](http://pve.splicedgene.com/raadpleging-versoepeling-coronamaatregelen); here a rebuilt [English Demo-version](https://burgerbegroting.tbm.tudelft.nl/participatory-value-evaluation-relaxation-corona-measures)), participants were presented with eight relaxation options which they could advise to the government;

1. Nursing and care homes allow visitors
2. Re-open businesses (other than contact professions and hospitality industry)
3. Re-open contact professions
4. Young people may come together in small groups
5. All restrictions lifted for people with immunity
6. All restrictions lifted in Northern provinces
7. Direct family members from other households can have social contact
8. Re-open hospitality and entertainment industry

More information about the relaxation options can be found through the [weblink](https://burgerbegroting.tbm.tudelft.nl/participatory-value-evaluation-relaxation-corona-measures). The order in which the options were presented was randomised across respondents. For each of these relaxation options, they received information regarding the option’s projected impact on the pressure on the health care system (which was expressed as the percentage in which the pressure on the health care system would increase due to the relaxation option). Moreover, for each option participants received information regarding its impact on increase of deaths among people younger than 70 years and older than 70 years, increase in the number of people with permanent physical injury, decrease in the number of people with permanent mental injury and the decrease in the number of households with long-term loss of income. For example, participants were shown that the relaxation option “re-open contact professions” would reduce the number of households that lose at least 15% of their income, but increase the number of deaths among people under the age of 70.

The constraint that participants faced in the PVE was the maximum capacity of the healthcare system in the sense that they were not able to recommend a bundle of relaxation options that in total resulted in a greater than 50% increase of the pressure on the healthcare system. Hence, they could only select a limited amount of relaxation options. Furthermore, participants were notified that the healthcare system could handle the pressure if it increased between 0% and 25%, that it would be overstretched if the pressure increased between 26% and 40%, and that it would be seriously overstretched if the pressure increased between 41% and 50%. After submitting their advice to the government, participants were asked to provide written motivations for their choices. Subsequently, they were asked which of the eight relaxation options should not be considered by the government and again they were asked to qualitatively underpin their choice. The main reason for including these open questions is that new arguments and ideas can emerge from the qualitative data and the government can learn about the arguments they can anticipate from those for and against specific relaxation options. Participants were also asked to answer various follow-up questions (e.g. gender, income, education and age) and they were also asked about the extent to which they themselves would experience impacts from each of the relaxation options they recommended to the government.

Each participant of the PVE faced one of 60 different profiles of relaxation options that varied on their impact levels and pressure to the healthcare system. These profiles were defined by following an experimental design process of three stages to avoid excessive correlation between impacts/pressure, and hence facilitate the statistical (econometric) analysis. First, a number of possible levels and pressure on the healthcare system were selected for each relaxation option, based on the information and feedback obtained from the early stages of the PVE design. The second stage consisted of constructing an initial candidate design of 60 profiles, by taking random impact and pressure levels from the first stage. The final stage consisted of iteratively replacing random impact or pressure levels in the candidate design, and evaluating on each iteration if the correlation between impacts/pressure is reduced. This process was repeated until a number of iterations without correlation reduction is reached, or after a time frame. For this particular design, we conducted the randomisation process for ten minutes, although we observed no further improvements after 3 minutes approximately. Appendix 1 provides a more detailed description of the possible impact and pressure levels, as well as the randomisation process and the correlation reduction criterion.

In the PVE, we made a substantial effort to ensure consequentiality, by (truthfully) informing respondents that the outcomes of this study would be shared with the Netherlands Institute of Public Health and Environment and high-ranking policymakers at relevant ministries. Consequentiality means that respondents must feel that their choices might have real -life consequences; the literature indicates that this substantially improves the reliability of the outcomes of preference elicitation studies^86-87^.

We carried out the PVE with two different samples. First, a randomly selected sample from the online Kantar Public panel, which was drawn to be representative of the Dutch population in terms of age and gender. Kantar Public approached members of their panel by e-mail to take part in our on-line survey and participants received a small monetary compensation. 3,358 respondents completed the experiment. The representative PVE was conducted to measure the preferences of ‘the average Dutch citizen’. A disadvantage of a ‘representative PVE’ is that only Dutch citizens that are part of the Kantar Public sample can participate. For this reason, we decided to open the PVE to the general public. A disadvantage of this ‘open PVE’ is that we, as researchers, have no control over which Dutch people participate and which do not. The results could be influenced by supporters or opponents of measures that mobilise many likeminded citizens. Hence, we carried out both a ‘representative PVE’ and an ‘open PVE’ because both have advantages and disadvantages. Our data collection effort was approved by the Ethics Board of the Delft University of Technology. Data was collected in the period 29 April – 4 May. Because our experiment was widely covered by the media, the number of participants was far higher than expected. As a result, the server could no longer cope with the volume and the PVE was offline on 30 April between 10.00 and 15.00. Eventually, 26,293 citizens participated in this ‘open PVE’. Appendix 2 presents socio-demographic characteristics of the participants.

### 4.3 Analysis of the data

The econometric framework to analyse people’s choices in a PVE is a Kuhn-Tucker type choice model based in the work of Bhat^88^, developed by Dekker et al.^26^ for PVE (henceforth, the MDCEV-PVE model), and adapted for this study. This framework is rooted in the consumer’s theory of microeconomics and relies on three key assumptions. First, it is assumed that an individual chooses the bundle of policy options that maximises their utility (i.e. satisfaction), subject to satisfying the resource constraint (in this case the limited capacity of the health sector). The second assumption is that part of the utility for each relaxation option depends on the impacts that are explicitly presented to individuals. For example, an individual may prefer relaxation options that reduce economic losses. Using the MDCEV-PVE model, the researcher can estimate so-called “taste parameters” to know the importance that individuals give to each impact on their choice of policy options. Additionally, the preferences for policy options can depend on other factors not associated with the impacts. The researcher can estimate so-called policy-specific constants to determine the benefits and costs individuals obtain from specific relaxation options, irrespective of the impacts that are explicitly communicated in the PVE. These policy-specific constants can also be complemented by including individual-specific variables to analyse sociodemographic differences in the preferences for relaxation options. Third, it is assumed that an individual can derive utility not only from (the impacts of) each relaxation option, but also from the resources that are not allocated. In the context of this PVE, individuals might want to advise against allocating the full capacity of the health care system because they do not want to overstretch the system.

We proceed to briefly formalize the MDCEV-PVE model used in this paper. Let *n* be an individual who faces *J* policy options and an amount of resources equal to *B*. When a policy *j* is chosen, it consumes a portion of *B* by an amount of *c*_*j*_. Following Dekker et al.^26^ specification of the individual’s utility function, the choice problem that individual *n* faces is given by:

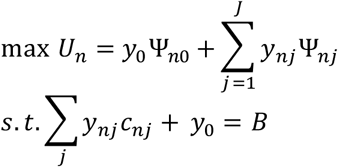

Where *y*_0_ is the amount of non-spent public resources, *y*_*nj*_ is a variable that takes value 1 if the individual chooses policy option *j* and zero otherwise, Ψ_*n*0_ is the utility provided by the non-spent resources, whereas Ψ_*nj*_ is the utility provided by the individual policy *j*. In the modelling, we assume that the utility for each policy option depends on the preferences for each known impact, as well as other factors apart from the impacts, encompassed in a policy-specific constant and sociodemographic characteristics. Therefore, we model the individual utility for policy options as Ψ_*nj*_ = exp(*δ*_*j*_ + Σ_*k*_ *β*_*k*_*x*_*njk*_ + Σ_*m*_ *θ*_*jm*_*z*_*nm*_ + *ε*_*nj*_), where *δ*_*j*_ is the specific constant for policy *j, β*_*k*_ is the taste parameter for impact *k, x*_*njk*_ is the level of impact *k* for policy *j, θ*_*jm*_ is a parameter that captures the extent that the sociodemographic characteristic *z*_*nm*_ affects the preferences for policy *j*, and *ε*_*nj*_ is an extreme-value type I stochastic term. The utility of non-spent resources is modelled in a similar form, by assuming Ψ_*n*0_ = exp(*δ*_0_ + *ε*_*n*0_). Dekker et al.^26^ provide an expression for the probability of choosing a bundle of policies under the MDCEV-PVE framework, allowing to estimate the model parameters using maximum likelihood.

The estimates of the MDCEV-PVE model can be used to determine the aggregate utility that a given bundle of policy options provides to society. Following Dekker et al. (2020), the aggregate utility of a given bundle of policies is given by:

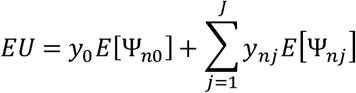

Where 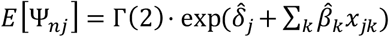 and 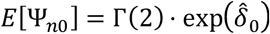. It is assumed that all individuals in society face the same levels of policy impacts. Thus, only a single level for each policy impact *x*_*jk*_ and *y*_0_ are considered for the computation of the aggregate utility. In general, these values are assumed to be the average value of each impact level and cost, for each policy option, or either the minimum or maximum levels when a sensitivity analysis of the aggregate utility is performed.

The aggregate utility function can be used to determine the bundle of policy options that maximizes the aggregate utility of society, provided that a policymaker has limited resources. Dekker et al.^26^ suggest a procedure to determine the optimal bundle by enumerating the aggregate utility of all possible combinations of policy options that satisfy a given resource limit and sorting them in descending order. The bundle with the highest aggregate utility is called the “optimal portfolio” of policy options.

Finally, the participants produced more than 100,000 written motivations for the choices they made in the PVE. A group of six annotators manually analysed randomly selected responses from 2,237 participants to provide an exhaustive list of arguments for and against each of the relaxation measures. One annotator experienced that saturation occurred after he had analysed the written motivations of 200 participants (no new arguments were added to the list of arguments), while another annotator had to review the responses of 500 participants to reach that point. The remaining annotators reached saturation between these two extremes. In a second round of analysis, three annotators counted the number of times that 600 respondents mentioned the arguments that were identified in the first round. The aim of this was to provide policymakers with information about the number of respondents who cited a specific argument. We could only include 600 respondents in this second round because the time between the start of our data collection and publication of our results for Dutch policymakers was very limited (29 April – 6 May).

## 5. Results

### 5.1 Descriptive results

The vast majority of participants supported a degree of relaxation of lockdown measures in the period 20 May – 20 July. We found little support for far-reaching relaxations that might cause the healthcare system to become heavily overloaded (higher than 41% increase in pressure on the health care system), but this varied across segments of the population. Figure 1 shows that men with high incomes and high education levels expressed a relatively strong preference for opening up (which would result in a relatively high pressure on the health care system). In contrast, older people on low incomes, who estimated that they themselves ran a high risk of becoming seriously ill from COVID-19, were relatively conservative in this regard. A further distinction is noticeable between the two survey groups. Participants in the representative PVE were significantly more cautious than participants in the open PVE in terms of their advice on relaxing lockdown measures. On average, participants in the ‘representative PVE’ recommended options resulting in a 28% increase in pressure on the healthcare system, while for those in the open PVE this was 32%. The percentage of participants advising against any relaxation whatsoever was much higher for the representative PVE than for the open PVE. This result suggests that citizens who participated in the open PVE were inclined to support a somewhat more extensive relaxation of lockdown measures than the average Dutch citizen (participants in the representative PVE).

**Figure 1:**
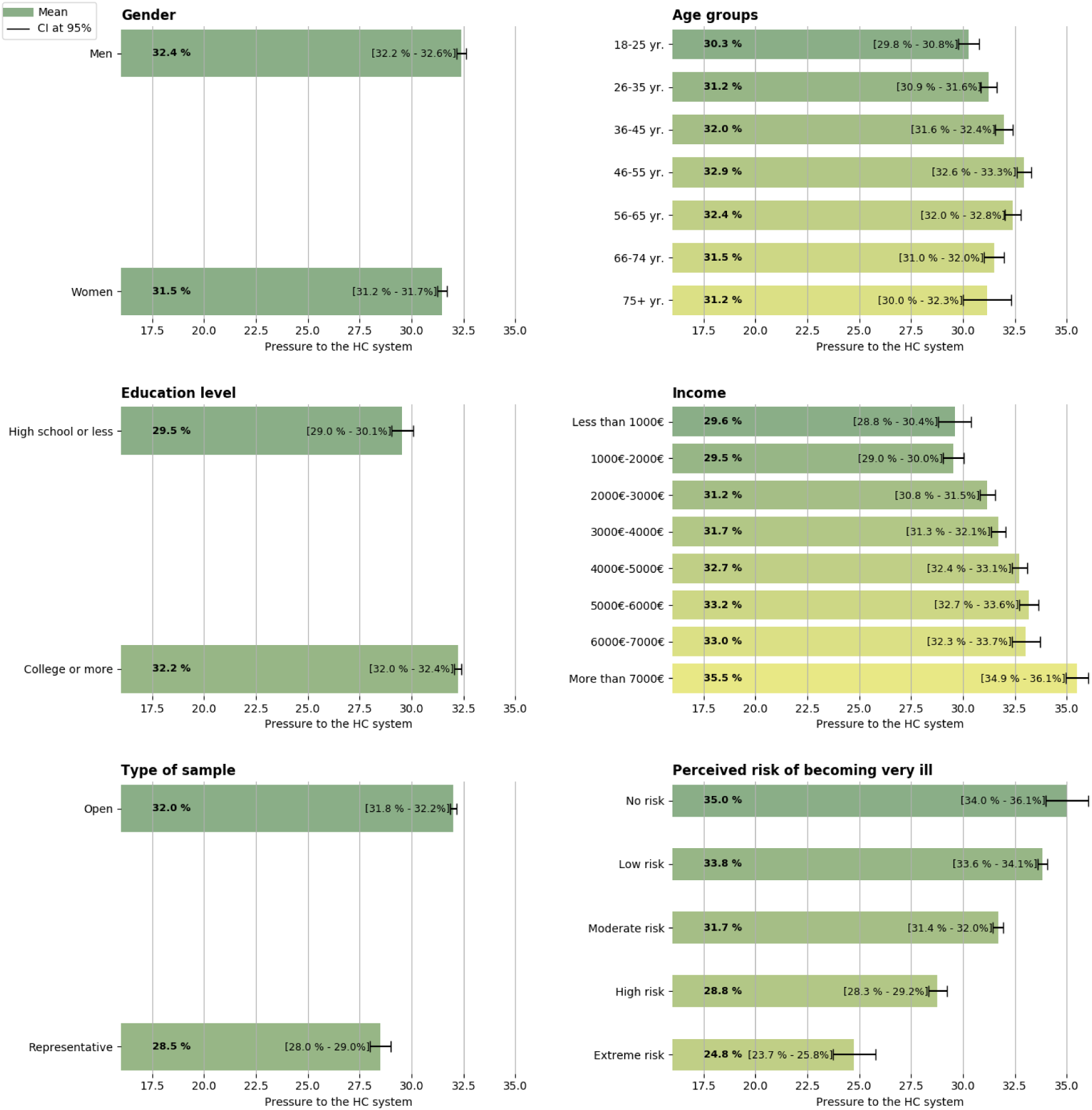
additional pressure of the health care system resulting from the recommended portfolio.

Figure 2 shows that in both the open PVE and the representative PVE participants most often recommended the option: “Re-open contact professions”. Figure 2 also shows that the strategy “Re-open hospitality and entertainment industry” was evaluated differently in the representative PVE and the open PVE. In the representative PVE 20% of the participants recommended this option and 45% discouraged this option, whilst in the open PVE the percentage of respondents who recommended this option was higher than the share of respondents opposing it. Moreover, Figure 2 shows participants divided about the desirability of the relaxation option ‘nursing and care homes should allow visitors’.

**Figure 2:**
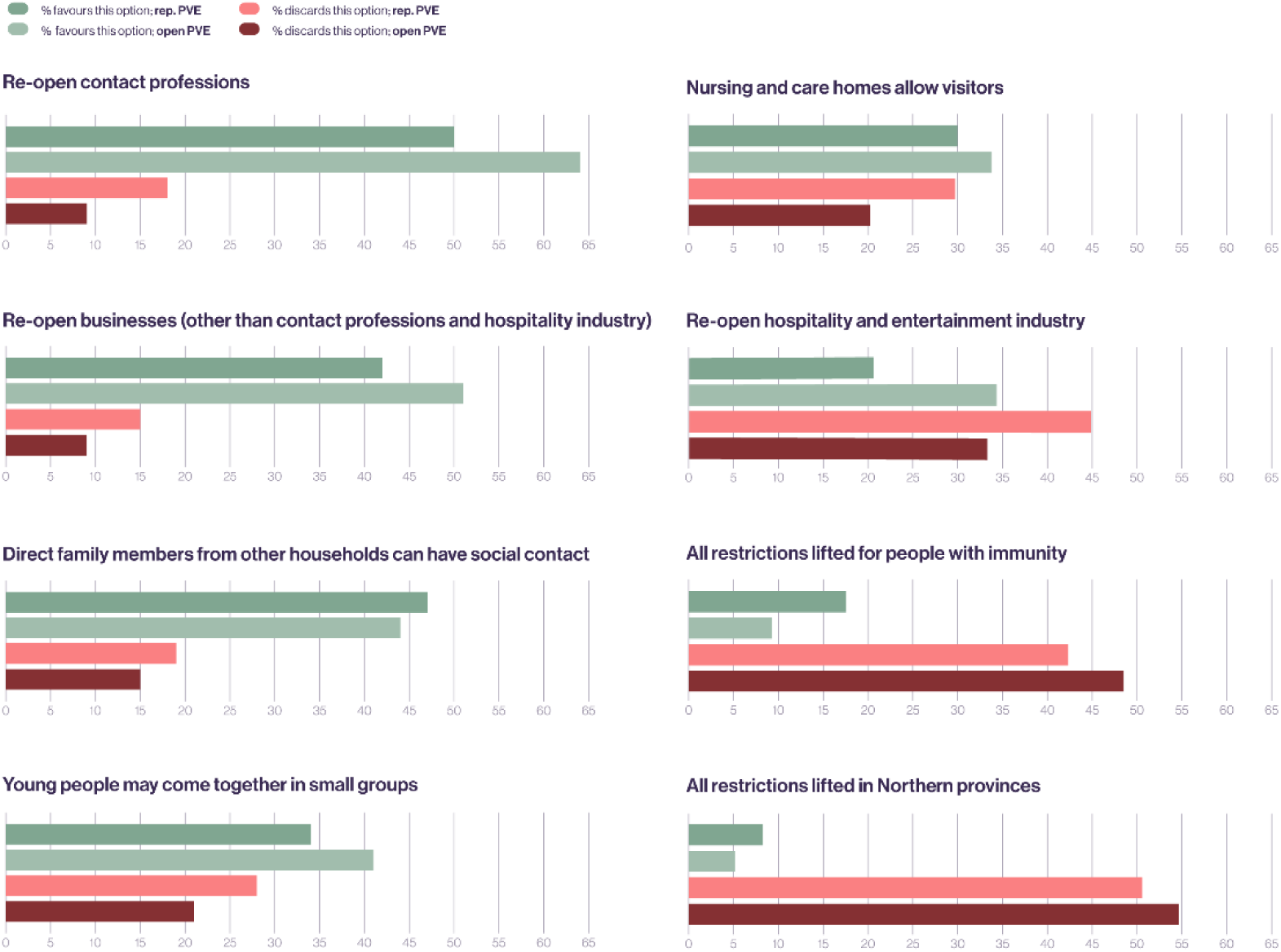
Percentage of respondents who recommended or opposed the eight relaxation measures.

One area of broad agreement was opposition to the relaxation of restrictions for specific groups of citizens. In both the representative PVE and the open PVE, the option “All restrictions lifted in Northern provinces” was least often advised, with “All restrictions lifted for people with immunity” not far behind. As seen in Figure 2, both options were rejected by more than 45% of the participants in the open PWE.

A normative objective in public participation is to secure distributional justice. The design of the PVE allowed citizens to consider the distributions of burdens and benefits of relaxing lockdown measures and enabled them to choose policy options from which they themselves would not benefit at all. To verify the extent to which participants choose relaxation policies that do (not) benefit themselves we asked them to indicate the impacts they predicted they would experience from each of the relaxation options they recommended. Table 2 shows that 71% of the respondents who recommended the relaxation option “Nursing and care homes allow visitors” would not personally experience any impacts from its implementation. 69% of the respondents would not expect to experience impacts from the relaxation option “Direct family members from other households can have social contact”. The written motivations (which we discuss more in detail in section 5.3) show that the interpretation of this result is ambiguous. On the one hand, there are respondents who choose this option for altruistic purposes. For instance, one respondent says: “I do not have any family, but I think that people who do have a family look forward to hold their loved ones”. On the other hand, many respondents said that the relaxation of this lockdown measure will not affect them as they already violated this rule.

**Table 2:** To what extent will lifting lockdown measures have an effect on your life?

|  | No effect | Small effect | Medium effect | Large effect | Very large effect |
| --- | --- | --- | --- | --- | --- |
| **Option 1** |  |  |  |  |  |
| Nursing and care homes allow visitors | 71% | 13% | 6% | 6% | 4% |
| **Option 2** |  |  |  |  |  |
| Re-open businesses (other than contact professions and hospitality industry) | 14% | 27% | 28% | 21% | 10% |
| **Option 3** |  |  |  |  |  |
| Re-open contact professions | 6% | 30% | 36% | 20% | 8% |
| **Option 4** |  |  |  |  |  |
| Young people may come together in small groups | 9% | 24% | 29% | 24% | 14% |
| **Option 5** |  |  |  |  |  |
| All restrictions lifted for people with immunity | 45% | 20% | 15% | 11% | 9% |
| **Option 6** |  |  |  |  |  |
| All restrictions lifted in Northern provinces | 51% | 19% | 15% | 9% | 6% |
| **Option 7** |  |  |  |  |  |
| Direct family members from other households can have social contact | 69% | 13% | 7% | 5% | 6% |
| **Option 8** |  |  |  |  |  |
| Re-open hospitality and entertainment industry | 14% | 19% | 26% | 25% | 16% |

### 5.2. Quantitative results

This section presents the estimation results of the MDCEV-PVE model under two specifications. In the first specification, we estimate a simple model that accounts for the effects of impacts through taste parameters as well as policy-specific constants. The second specification includes sociodemographic variables for each relaxation option to uncover differences between different groups of individuals in terms of their preferences over certain relaxation options. We then provide the optimal portfolio of relaxation options for the first specification. All results provided in this section were calculated using the full available sample (i.e. combining responses from the open sample and the representative sample). Appendix 3 provides the estimation results of the first specification of the MDCEV-PVE model for each sample separately.

#### 5.2.1. MDCEV-PVE model estimates

Table 3 summarises the MDCEV-PVE estimates for the model without sociodemographic variables, henceforth referred to as the “simple model”. The first set of estimates are the taste parameters. All estimates are statistically significant, except for the taste parameter associated with reductions in permanent mental injuries. The sign of the taste parameters indicates whether an increase in the associated impact makes a relaxation option more (un)attractive. Thus, any additional deaths and (permanent) physical injuries resulting from COVID-19 negatively impact the attractiveness of a relaxation option, while a reduction in the number of households experiencing income loss of greater than 15% increases that attractiveness. Using the taste parameters, it is also possible to establish the relative importance of the different impacts in defining the desirability of relaxation options. For instance, we can infer from the results that citizens consider a reduction of 100 deaths of persons below the age of 70 years and the reduction of 168 deaths of citizens older than 70 years (−0.8486 / -0.5084) equally attractive (in that they provide the same utility).

**Table 3:** MDCEV model estimates.

|  | Estimate | (Std. Err.) |
| --- | --- | --- |
| **Policy-specific constants:** |  |  |
| 1: Nursing and care homes allow visitors | 2.6948*** | (0.0273) |
| 2: Re-open businesses (other than contact professions and hospitality industry) | 2.6187*** | (0.0208) |
| 3: Re-open contact professions | 3.1906*** | (0.0243) |
| 4: Young people may come together in small groups | 1.8544*** | (0.0127) |
| 5: All restrictions lifted for people with immunity | 1.6231*** | (0.0200) |
| 6: All restrictions lifted in Northern provinces | 1.6617*** | (0.0314) |
| 7: Direct family members from other households can have social contact | 2.5117*** | (0.0278) |
| 8: Re-open hospitality and entertainment industry | 2.7032*** | (0.0327) |
| **Taste parameters:** |  |  |
| Additional 10.000 deaths of people of +70 years | -0.5084*** | (0.0802) |
| Additional 10.000 deaths of people of less than 70 years | -0.8486*** | (0.1582) |
| Additional 10.000 people with permanent physical injury | -0.1082*** | (0.0155) |
| Minus 10.000 people with permanent mental injury | 0.0006 | (0.0033) |
| Minus 10.000 households that have lost 15% of income | 0.0076*** | (0.0022) |
| Observations | 29651 |  |
| Log-likelihood | -144957.5115 |  |
| AIC | 289889.0230 |  |
| BIC | 289781.1588 |  |
| **Statistical significance:** ***p < 0.001, **p < 0.01, *p < 0.05 |  |  |

The second set of estimates correspond to the policy-specific constants. A higher value of these estimates reflects a stronger preference for the associated relaxation options irrespective of the impacts for which we estimated taste parameters.

Table 4 summarizes the estimates of an MDCEV-PVE model which includes a set of sociodemographic variables for each relaxation option. We included a variable to identify potential differences in the preferences of men and women, a variable to identify the extent to which the preferences of the youngest (19 to 25 years old) and oldest (above 65 years old) citizens differ from those in the middle age groups and a variable to analyse whether people with a high education level have different preferences than those with a lower education level. Finally, we analysed whether residents of the Northern provinces have stronger preferences for lifting all restrictions in their own region.

**Table 4:** Estimation results of MDCEV-PVE model (with covariates)

|  | Nursing homes | Businesses | Contact professions | Young people | People with immunity | Northern provinces | Direct family members | Hospitality industry |
| --- | --- | --- | --- | --- | --- | --- | --- | --- |
| **Parameters specific to each relaxation option** |  |  |  |  |  |  |  |  |
| Constant | 2.8651***<br>(0.0384) | 2.1238***<br>(0.0332) | 3.0668***<br>(0.0332) | 1.6067***<br>(0.0279) | 1.6348***<br>(0.0350) | 1.5363***<br>(0.0450) | 2.6065***<br>(0.0385) | 2.4668***<br>(0.0404) |
| Is Male | -0.4537***<br>(0.0233) | 0.4094***<br>(0.0244) | 0.1953***<br>(0.0263) | 0.0576*<br>(0.0233) | 0.1414***<br>(0.0259) | 0.0884**<br>(0.0299) | -0.1178***<br>(0.0236) | 0.2213***<br>(0.0231) |
| Is above 65 years old | 0.4480***<br>(0.0335) | -0.0151<br>(0.0348) | -0.2021***<br>(0.0370) | 0.0130<br>(0.0331) | 0.0552<br>(0.0358) | 0.2171***<br>(0.0394) | -0.0591<br>(0.0339) | -0.5659***<br>(0.0334) |
| Is between 19 and 25 years old | -0.3941***<br>(0.0399) | 0.0100<br>(0.0410) | -0.2833***<br>(0.0425) | 0.4105***<br>(0.0409) | -0.1289**<br>(0.0452) | -0.2468***<br>(0.0558) | -0.0612<br>(0.0395) | 0.1659***<br>(0.0391) |
| Has college degree (HBO or university) | 0.0872**<br>(0.0271) | 0.4254***<br>(0.0276) | 0.1660***<br>(0.0300) | 0.2649***<br>(0.0271) | -0.1252***<br>(0.0295) | -0.0448<br>(0.0339) | -0.0321<br>(0.0277) | 0.2359***<br>(0.0269) |
| Lives in a Northern province |  |  |  |  |  | 0.8371***<br>(0.0418) |  |  |
| **Taste parameters (common among all relaxation options)** |  |  |  |  |  |  |  |  |
| Additional 10.000 deaths of people of +70 years |  |  |  |  | -0.5955***<br>(0.0956) |  |  |  |
| Additional 10.000 deaths of people of less than 70 years |  |  |  |  | -0.8803***<br>(0.2022) |  |  |  |
| Additional 10.000 people with permanent physical injury |  |  |  |  | -0.1148***<br>(0.0165) |  |  |  |
| Minus 10.000 people with permanent mental injury |  |  |  |  | 0.0034<br>(0.0036) |  |  |  |
| Minus 10.000 households that have lost 15% of income |  |  |  |  | 0.0091***<br>(0.0024) |  |  |  |
| Observations |  |  |  |  | 24004.0000 |  |  |  |
| Log-likelihood |  |  |  |  | -115791.0719 |  |  |  |
| AIC |  |  |  |  | 231490.1437 |  |  |  |
| BIC |  |  |  |  | 231118.1888 |  |  |  |

**Note:** Standard errors in parenthesis. **Statistical significance:** \*\*\* p < 0.001, \*\* p < 0.01, \* p < 0.05.

Our results support the existence of varying preferences for relaxation options among different sociodemographic groups. We observe that the estimated parameters associated with sociodemographic variables are in general statistically significant. The sign of these parameters indicates whether individuals who belong to the sociodemographic group perceive the relaxation option as more (un)attractive.

We can illustrate this with a few examples from the results. In terms of gender differences, men perceive allowing visitors in nursing homes as less attractive than women do; at the same time, however, men are more positive about re-opening contact professions. With respect to age, people above 65 years old are most supportive of allowing visitors in nursing homes, while those between the ages of 19 and 25 are more receptive to a re-opening of the hospitality industry than are other age groups. In terms of education level, Dutch citizens with a higher level of education perceive re-opening the hospitality industry as more attractive than people with other educational backgrounds. Finally, residents of the Northern provinces perceive lifting restrictions in that region as more attractive than inhabitants of other provinces. One of the results that stands out is that the estimated parameters for the option “re-open contact professions” are consistently small regardless of socioeconomic grouping, while the policy-specific constant is the highest out of any option. This indicates a broad base of support throughout Dutch society. We also estimated an MDCEV-PVE simple model using a sample of residents of the Northern provinces and report the results in Appendix 4. Although citizens living in this region have a relatively positive view of the strategy which entails lifting the corona measures in the Northern provinces, this strategy is not included in the optimal portfolio.

#### 5.2.2 Optimal portfolios of relaxation options

Using the estimates of Table 3, we computed the optimal portfolio of relaxation options which respects the budget constraint of a maximum increase of the pressure to the healthcare system of 50%. This optimal portfolio is determined under the assumption that all individuals in society face the same impact levels and pressure on the healthcare system. We have taken these values from the average impact levels and pressure presented in the experiment (see Appendix 5). We include two additional scenarios for the purpose of sensitivity analysis. The first scenario is a pessimistic case, under the assumption that all individuals in society face the maximum levels of pressure to the healthcare system, the maximum levels of the impacts that have a negative taste parameter estimate, and the minimum levels of the impacts that have a positive taste parameter estimate. The second scenario is an optimistic case, in which all individuals in society face the minimum levels of pressure, the minimum levels of impacts with negative taste parameter estimates, and the maximum levels of impacts with positive taste parameter estimates. More information on the impact levels and pressure used to compute the optimal portfolios in these sensitivity analyses can also be found in Appendix 5.

Table 5 lists the optimal portfolio under each of the three scenarios. The optimal portfolio given an average-level scenario suggests that the most preferred bundle of relaxation options is to re-open contact professions re-open businesses (except the hospitality industry) and to allow social contact again between families. This bundle imposes an increase of the pressure to the healthcare system of 32%, still leaving a substantial amount of pressure without allocation. Under a pessimistic scenario, only allowing contact professions to re-open is included in the optimal portfolio, with a pressure to the healthcare system of 15%. Finally, under an optimistic scenario, five out of eight relaxation options are part of the optimal portfolio, excluding re-opening the hospitality industry, lifting restrictions for individuals with immunity and lifting restrictions for the Northern provinces. Such bundle of relaxation policies results in an increase in the pressure to the healthcare system of 34%.

**Table 5:**
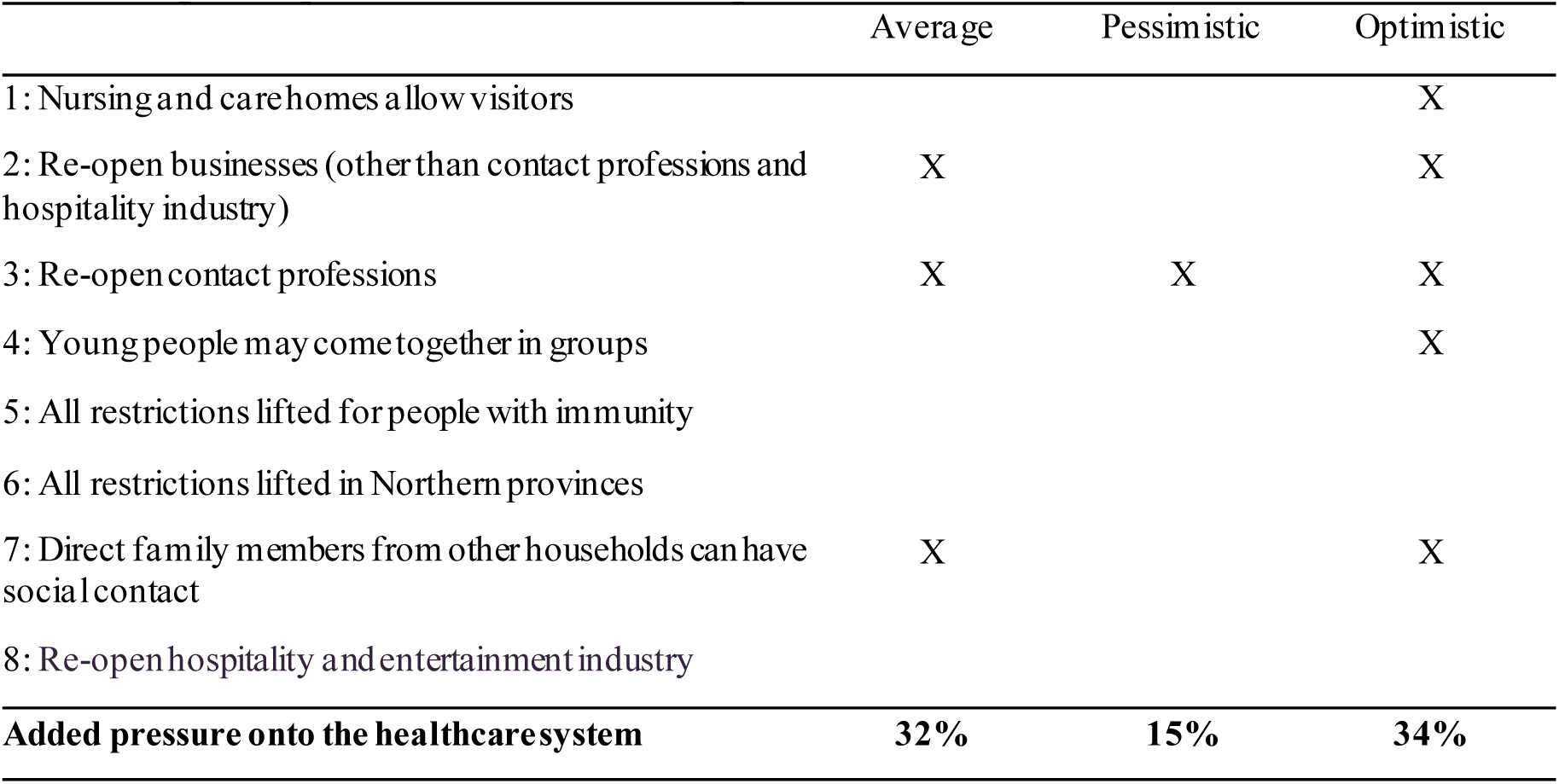
Optimal portfolios of relaxation options.

|  | Average | Pessimistic | Optimistic |
| --- | --- | --- | --- |
| 1: Nursing and carehomes allow visitors |  |  | X |
| 2: Re-open businesses (other than contact professions and hospitality industry) | X |  | X |
| 3: Re-open contact professions | X | X | X |
| 4: Young people may come together in groups |  |  | X |
| 5: All restrictions lifted for people with immunity |  |  |  |
| 6: All restrictions lifted in Northern provinces |  |  |  |
| 7: Direct family members from other households can have social contact | X |  | X |
| 8: Re-open hospitality and entertainment industry |  |  |  |
| **Added pressure onto the healthcare system** | **32%** | **15%** | **34%** |

### 5.3 Qualitative results

Analysis of the written motivations of 2,237 randomly selected participants on why they preferred some relaxation options over others revealed four insights. First, it shed light on the arguments and concerns of proponents and opponents of each option; many of these had not come to our attention during our analysis of media content and the conversations that we had with policymakers when designing the PVE. We will summarize all arguments in Tables 6-13 and also show how many respondents of the 600 respondents from whom written motivations were analysed in the second round cited a certain argument. The second insight relates to the underlying principles that are at stake in relaxing lockdown measures. For example, participants consider it important that the relaxation leads to “unity” rather than “division”. These principles seem to play a large role in the explanation of why certain relaxation options are not favoured by Dutch citizens (e.g. lifting restrictions for the Northern provinces or for Dutch people who are immune to COVID-19). The third insight relates to how Dutch citizens condition their preferences. Without being specifically asked, a large number of participants conditioned their relaxation preferences to, amongst other things, increased safety measures. These conditions also revealed ideas, how to solve dilemmas of relaxation options and improve the effectiveness of relaxation options. Finally, the fourth insight was hearing explicitly from participants that they had evaluated relaxation options in relation to each other. This supports the use of PVE as a preference elicitation technique over alternatives such as CV and DCE, as it is a key advantage of the former.

**Table 6:**
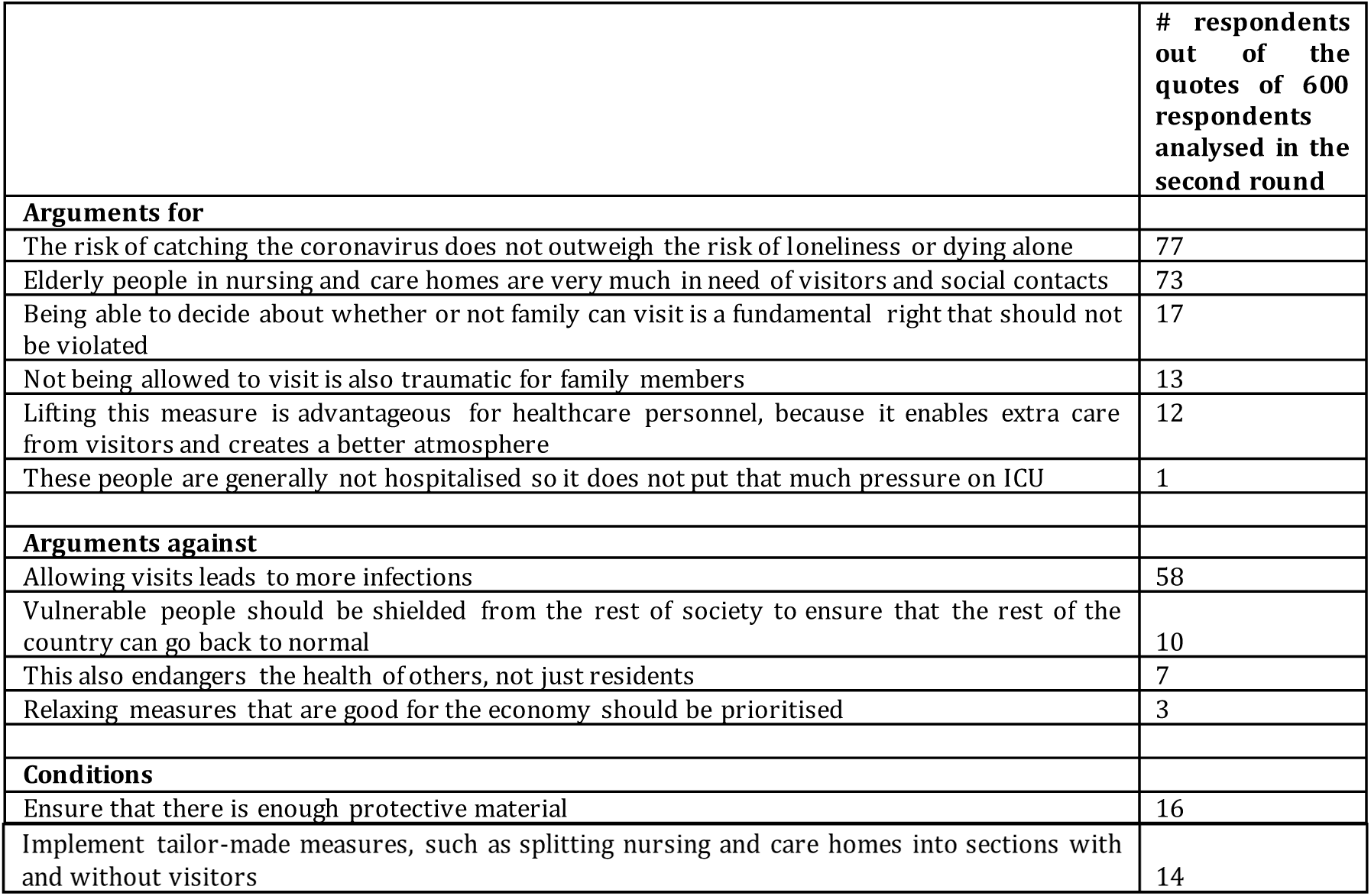
Nursing and care homes allow visitors: arguments for, arguments against and conditions.

**Table 7:** re-open businesses other than contact professions and hospitality industry: arguments for, arguments against and conditions.

|  | # respondents out of the quotes of 600 respondents analysed in the second round |
| --- | --- |
| **Arguments for** |  |
| This option prevents substantial damage to the economy | 187 |
| Being able to work again has a positive effect on people's well-being and mental health | 59 |
| The impact on the number of infections will not be high | 13 |
| If we don't get the economy out of the muck quickly, we won't be able to pay for our expensive health care in the future. The money that is needed to finance the health care sector needs to be earned somewhere | 9 |
| When developed countries close their economy this will amplify poverty in developing countries | 1 |
| **Arguments against** |  |
| This measure substantially increases the risk of infections as businesses bring large groups of people together and will also result in greater movement of persons throughout the Netherlands | 27 |
| Working from home is not so bad | 16 |
| **Conditions** |  |
| Only when social distancing and/or isolated workplaces can be guaranteed at the office | 67 |
| There should also be an option for high-risk individuals to work from home | 22 |

**Table 8:**
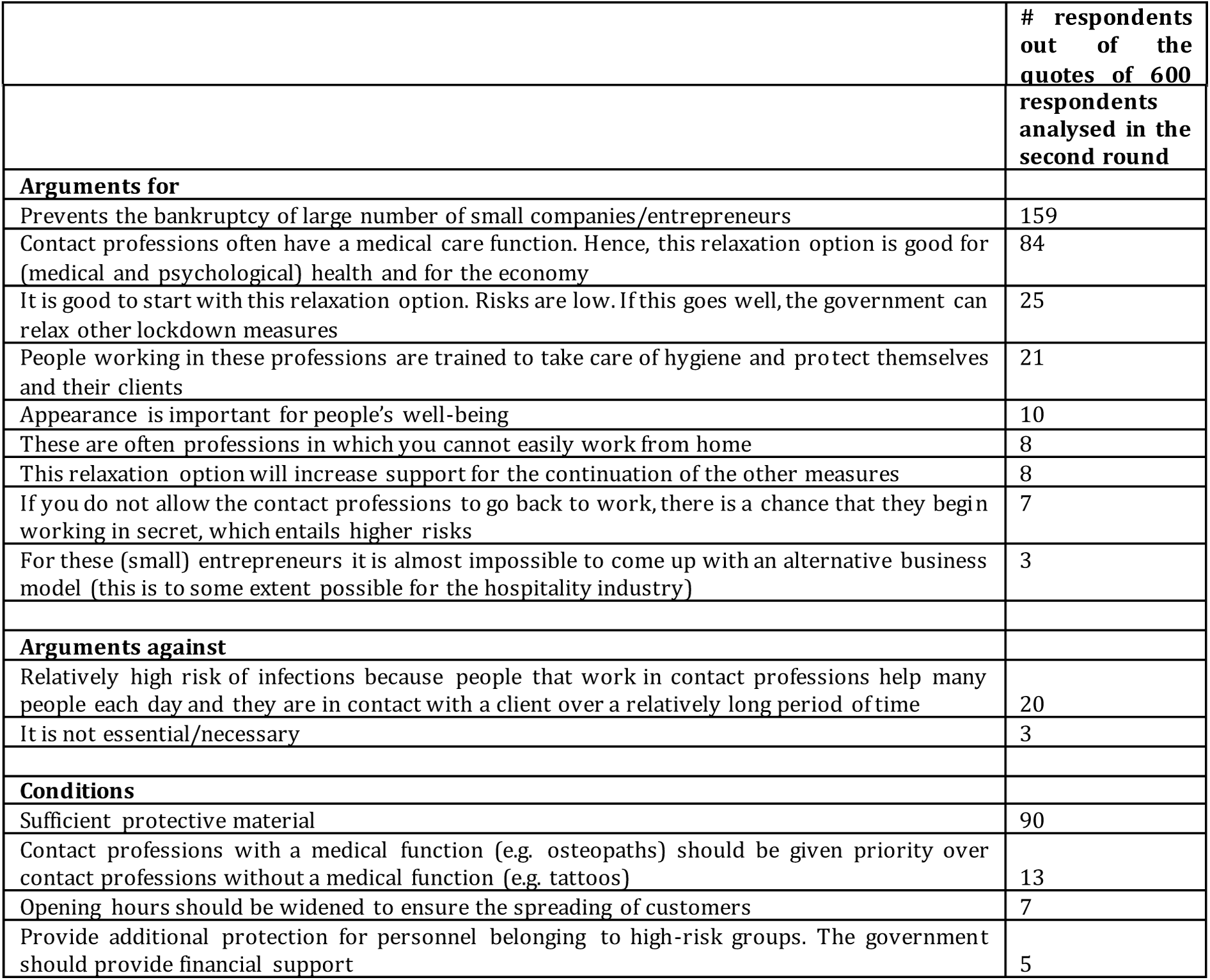
Re-open contact professions: arguments for, arguments against and conditions.

|  | # respondents out of the quotes of 600 |
| --- | --- |

|  | respondents analysed in the second round |
| --- | --- |
| **Arguments for** |  |
| Prevents the bankruptcy of large number of small companies/entrepreneurs | 159 |
| Contact professions often have a medical care function. Hence, this relaxation option is good for (medical and psychological) health and for the economy | 84 |
| It is good to start with this relaxation option. Risks are low. If this goes well, the government can relax other lockdown measures | 25 |
| People working in these professions are trained to take care of hygiene and protect themselves and their clients | 21 |
| Appearance is important for people's well-being | 10 |
| These are often professions in which you cannot easily work from home | 8 |
| This relaxation option will increase support for the continuation of the other measures | 8 |
| If you do not allow the contact professions to go back to work, there is a chance that they begin working in secret, which entails higher risks | 7 |
| For these (small) entrepreneurs it is almost impossible to come up with an alternative business model (this is to some extent possible for the hospitality industry) | 3 |
| **Arguments against** |  |
| Relatively high risk of infections because people that work in contact professions help many people each day and they are in contact with a client over a relatively long period of time | 20 |
| It is not essential/necessary | 3 |
| **Conditions** |  |
| Sufficient protective material | 90 |
| Contact professions with a medical function (e.g. osteopaths) should be given priority over contact professions without a medical function (e.g. tattoos) | 13 |
| Opening hours should be widened to ensure the spreading of customers | 7 |
| Provide additional protection for personnel belonging to high-risk groups. The government should provide financial support | 5 |

**Table 9:** Young people may come together again in small groups: arguments for, arguments against and conditions.

|  | # respondents out of the quotes of 600 respondents analysed in the second round |
| --- | --- |
| **Arguments for** |  |
| Young people play a minor role in the spread of the virus and their risk of getting sick is low | 136 |
| Social contact is relatively important for young people (to develop themselves) | 61 |
| For young people it is difficult not to violate the rules | 49 |
| Reduction of problematic psychological symptoms | 18 |
| Reduces the pressure on parents | 17 |
| Possibility to build up herd immunity | 10 |
| Increases support among young people for other lockdown measures | 5 |
| **Arguments against** |  |
| Constitutes age discrimination which results in a dichotomy in society | 27 |
| Measures are difficult to enforce. Young people will also get in contact with other people | 23 |
| **Conditions** |  |
| Young people should maintain 1.5m distance from those outside that group | 20 |

**Table 10:** All restrictions lifted in the Northern provinces: arguments for, arguments against and conditions.

|  | # respondents out of the quotes of 600 respondents analysed in the second round |
| --- | --- |
| **Arguments for** |  |
| Low risk of transmission in these provinces. The impact of relaxation measures can be monitored relatively easily | 15 |
| Impact of relaxing lockdown measures can be monitored and this provides useful information for future decisions on relaxing lockdown measures | 9 |
| Boosts the economy in the North of the Netherlands | 7 |
| **Arguments against** |  |
| Practically unfeasible because this is almost impossible to enforce. People will go to the North for entertainment and bring infections to these provinces | 122 |
| Solidarity will be undermined and this will not benefit the Netherlands as a whole | 113 |
| **Conditions** |  |
| Enforceability of this measure should be guaranteed | 3 |
| Measures should be relaxed in small steps | 1 |

**Table 11:** All restrictions lifted for people with immunity: arguments for, arguments against and conditions.

|  | # respondents out of the quotes of 600 respondents analysed in the second round |
| --- | --- |
| **Arguments for** |  |
| These people pose no danger to their environment | 16 |
| These people can keep society and the economy going again | 10 |
| It is pointless to demand solidarity from these people if they are already immune. Doing so will lead to fierce protests | 9 |
| **Arguments against** |  |
| Tests for immunity are not foolproof, and this increases the risk of new infections | 121 |
| Creates a dichotomy in society. People who are not immune can get annoyed by the behaviour of those who are allowed to resume normal life | 70 |
| Difficult to enforce | 60 |
| Potential confusion as immunity is not outwardly apparent | 18 |
| **Conditions** |  |
| Only consider this option when you are 100% sure that immunity can be measured | 1 |

**Table 12:** Direct family members from other households can have social contact: arguments for, arguments against and conditions.

|  | # respondents out of the quotes of 600 respondents analysed in the second round |
| --- | --- |
| **Arguments for** |  |
| Improves the well-being of many Dutch people. Contact with family is important in times of crisis, and can alleviate psychological harm. Hence, in the longer term, this can reduce the need for mental care caused by psychological distress | 123 |
| People will behave responsibly to ensure that they do not infect their loved ones. Family members keep each other informed about their health | 46 |
| People already violate this rule so this relaxation option brings the rules more in sync with reality | 41 |
| This allows contact with only a small number of people ('social bubble') which has a relatively small impact on the risk of large-scale transmission of COVID-19 | 26 |
| This relaxation option ensures that citizens will comply with the lockdown measures. It provides positive energy and optimism over the future | 8 |
| Grandparents can take care of their grandchildren which reduces pressure on families | 5 |
| **Arguments against** |  |
| This substantially increases the risk of infections | 16 |
| This measure is difficult to enforce | 10 |
| Focusing only on (direct) family is too limited. My friends are more important to me than my family | 7 |
| Measures that have an impact on the economy should be prioritised | 3 |
| **Conditions** |  |
| Ensure that this rule is only applicable to direct family members | 6 |

**Table 13:** Re-open hospitality and entertainment industry: arguments for, arguments against and conditions.

|  | # respondents out of the quotes of 600 respondents analysed in the second round |
| --- | --- |
| **Arguments for** |  |
| This is good for our economy and business | 106 |
| It is good for people's well-being | 83 |
| This relaxation option will increase support for the continuation of the other measures | 7 |
| It is enforceable | 7 |
| People can take responsibility for themselves by staying away if they wish | 7 |
| We should preserve our cultural heritage and cannot risk bankruptcies in the cultural sector | 4 |
| Keeping these businesses closed is too big of a sacrifice for young people | 3 |
| In this way, we can build up herd immunity | 1 |
| If the hospitality industry is not re-opened people will do other things to relax which is also risky | 1 |
| **Arguments against** |  |
| Risk of too many people gathering together, which helps to spread the virus | 83 |
| It is not necessary at the moment | 22 |
| When alcohol is consumed, people are more likely to underestimate risks and are less likely to comply with distancing measures | 11 |
| Opening up the hospitality and entertainment sectors should only be considered in the next phase if it appears that other adjustments have worked | 10 |
| Hospitality industry has a bad impact on society. Please keep it closed | 1 |
| **Conditions** |  |
| There are many options for measures to be taken in hospitality and entertainment (including reducing alcohol consumption). Rely on the sector's creativity and sense of responsibility. | 40 |
| It is important to differentiate between different sectors (e.g. bars closed, museums open) | 14 |
| Re-open hospitality industry but restrict opening hours | 1 |

#### 5.3.1 Nursing and care homes allow visitors

Many participants who recommended this option argue that the quality of life of older people and those in their final stages of life is more important than increasing their life expectancy. In sections 5.1. and 5.2 we already showed that participants were divided about the attractiveness of this relaxation option and the written motivations also reflect strong differences in opinion among respondents regarding the desirability of this strategy. On the one side, many respondents refer to fundamental rights when arguing that inhabitants of nursing and care homes should be able to decide for themselves whether they want to allow visitors. On the other hand, some respondents who disfavour this option argue that old and vulnerable people should be shielded from the rest of society to ensure that the rest of the country can go back to normal. Moreover, various respondents argued that relaxation options that positively impact the economy should be prioritised, which suggests that they evaluated this relaxation option in relation to the others.

#### 5.3.2 Re-open businesses other than contact professions and hospitality industry

Many participants indicated that they selected this option because of the benefits for the economy, which is an argument that was anticipated a priori based on our conversations with policymakers. Nevertheless, the relatively large number of people who revealed generally positive attitudes over working from home was quite surprising. In meetings with policy makers and the media we did not encounter the argument that opening-up the economy in developed countries will have positive impacts on people living in developing countries.

#### 5.3.3 Re-open contact professions

This relaxation option was the most chosen option by respondents and section 5.2 shows that there was also widespread support for this option among participants from different socio-demographic groups. Table 8 shows that participants provided a range of arguments as to why this option should be prioritised by the government. From the written motivations it could be inferred that many participants in the PVE sympathised with preventing the bankruptcy of a large number of (generally small) businesses. Another argument cited was that contact professions should re-open because, unlike the hospitality industry, they lack alternative sources of income. This was an argument that was not raised in the media content that we analysed, nor in the conversations that we had with policymakers in the design stage of the PVE. The fact that respondents explicitly made a comparison with the circumstances in the hospitality industry provides evidence that participants valued relaxation options in relation to each other rather than separately. Various respondents argued that contact professions with a medical purpose should be prioritised.

#### 5.3.4 Young people may come together again in small groups

Under this option, young people would still be required to respect the 1.5 -metre distance rule when they meet older people. Supporters cited its relatively small effect on the spread of the virus, the low risk for young people and its positive effects for young people, while detractors saw problems around enforcement of this rule and its being seen as a form of age discrimination.

#### 5.3.5 All restrictions lifted in the Northern provinces

Sections 5.1 and 5.2 reveal that there is little support among Dutch citizens for policy options that relax restrictions for one specific group of citizens. Many participants find it very important that the relaxation of lockdown measures leads to “unity” and not to “division”. They are afraid that the unity among Dutch people that currently exists – along with the support for corona-related government policies – will be lost if and when the Cabinet chooses to lift restrictions for a specific group of Dutch people (e.g. the North of the Netherlands, Dutch people who are immune to COVID-19). Below are several quotes that illustrate this point:

*“By making a distinction between people who are immune and people who may still be infected, you create a very strange dividing line between two groups in the population. The same with all the restrictions lifted in the Northern provinces. It’s either the whole of the Netherlands without restrictions, or not. Making divisions between occupations or parts of daily life (such as hospitality vs. contact professions) to lift restrictions is about smaller steps and is easier to understand than exempting a whole part of the Netherlands”*.

*“We have to overcome this crisis together, so it is not wise to create divisions”*.

*“There should be no difference between people. We live in one country and all have to follow the same rules. We are all Dutch and that means equal treatment”*.

*“We are a country of 17 million people, who should be treated equally. We fight for equality and against racism so you should not make a distinction between people that live in different parts of the country*.*”*

#### 5.3.6 All restrictions lifted for people with immunity

The relaxation option “For people who are immune, all restrictions are lifted” can also count on little support from the Dutch population. Table 11 shows that people have various concerns about this option and also cite that they oppose this option because it might lead to a dichotomy in society.

#### 5.3.7 Direct family members from other households can have social contact

Some of the written motivations provided by respondents who advised this relaxation option were new and unexpected. For instance, various respondents argued that they selected this option because, in their view, this will increase compliance with other lockdown measures as it provides positive energy and optimism. Moreover, it is noteworthy that many respondents supported this option because, in their view, many people (sometimes including respondents themselves) already violated this lockdown measure. On the other hand, various participants disfavoured this option as they argued that, for them, seeing friends was more important than social contact with family.

#### 5.3.8 Re-open hospitality and entertainment industry

Participants argued that opening up hospitality and entertainment is not only good for the economy and business, but they also considered it important for the well-being of the Dutch. That said, many participants were also concerned that this relaxation option would result in increased infections, particularly in situations where the consumption of alcohol had the potential to change perceived risks for individuals. Many participants argued that this relaxation option is less urgent than other options. For instance, one respondent argued that “nursing and care homes allow visitors” should be prioritised because the situation in nursing and care homes is much more poignant. Finally, some participants argued that the risk that this relaxation option contributes to new outbreaks of COVID-19 is relatively high and for this reason they think that this option should only be considered after other options had turned out to be successful.

### 5.4 The merits of the PVE as perceived by participants

The draft results of our study were shared on 4 May with the Ministry of Health and the Dutch National Institute for Public Health. The latter, in turn, chairs the central Outbreak Management Team which advises the government on COVID-19 policies. The final results were shared on 6 May. As their involvement and collaboration in the research showed, those experts were open to and cognisant of concerns and priorities from the public. We do not know whether and how our results affected political decisions on the relaxation of lockdown measures, but it is noteworthy that the Dutch government decided on 6 May to start with the relaxation of lockdown measures for contact professions which was in line with our result that re-opening contact professions would have broad support in society. Another example of the way that political decisions overlapped with our results is that the Dutch government, unlike other countries such as Germany, adopted a central approach in terms of imposing and relaxing lockdown measures without differentiating between regions.

In section 3, we proposed several hypotheses regarding the strengths of PVE. These related to enabling citizens to participate in multi-dimensional policy issues (normative rationale for participation) and letting citizens experience intricate government dilemmas so as to improve their understanding of relevant trade-offs and potentially improve future compliance (instrumental rationale for participation). Moreover, we discussed that a potential weakness of PVE is that the quality of preferences that people express is probably lower than preferences that they express after deliberation (which is where mini-publics have an advantage).

To explore the extent to which the hypothesised strengths and weaknesses were actually realised, we evaluated how participants experienced their participation in the PVE through asking them to respond to several propositions (see Figure 3) and we asked open questions to reflect on the strengths and weaknesses of the method. Table 14 provides an overview of the number of respondents that cited a certain strength out of the 600 respondents for whom written motivations were analysed in the second round of analysis.

**Table 14:** number of times that perceived strengths of the PVE method were cited.

| Perceived strength | # respondents out of the quotes of 600 respondents analysed in the second round |
| --- | --- |
| The survey was very clear (clear instruction video and background information) | 88 |
| **Substantive rationale for participation** |  |
| This is an informed advice to the government based on insights regarding the consequences of your advice | 76 |
| Provides lot's opportunities to explain my advises and to add nuances | 49 |
| The constraint forces participants to make a choice (not possible to just choose everything) | 10 |
| The government gets an impression of citizens' preferences regarding this topic | 4 |
| **Normative rationale for participation** |  |
| Positive that the government consults its citizens | 52 |
| I had the feeling that my opinion counted | 4 |
| Positive that the consultation was accessible for all citizens. | 2 |
| Allowed me to provide a contribution to fighting the COVID-19 crisis | 1 |
| **Instrumental rationale for participation** |  |
| Raised my awareness regarding (consequences of) relaxation options | 77 |
| Improves transparency regarding the dilemmas the government faces | 34 |
| Encourages me to reflect on my own opinions | 7 |
| Improves understanding and support for final decisions on relaxation of lockdown measures | 5 |

**Figure 3:**
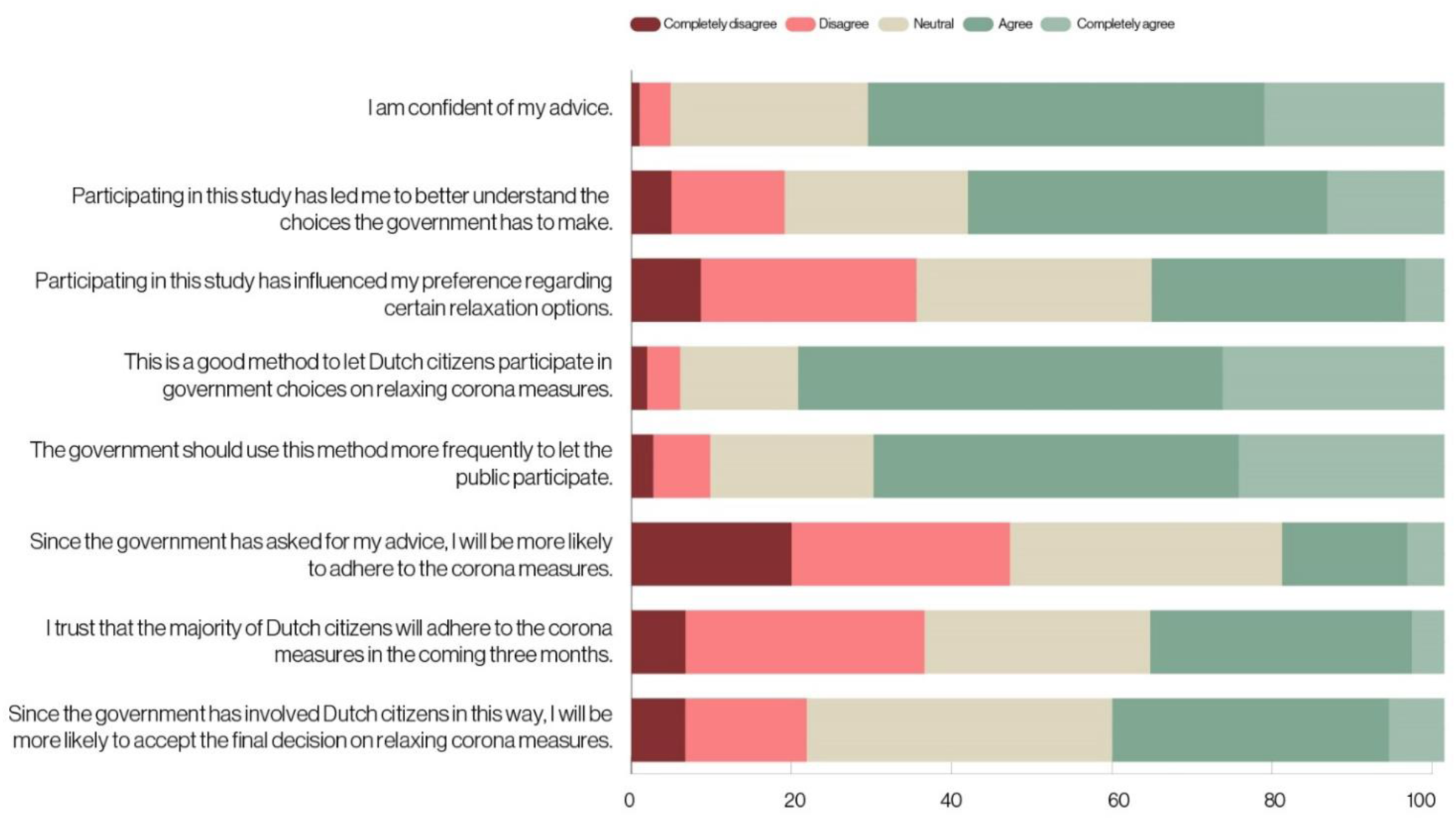
experiences of participants and their likelihood of adherence/acceptance of measures.

A first perceived strength of PVE is that putting large groups of people in the shoes of a policymaker might raise their awareness of intricate government dilemmas. Figure 3 shows that around 60% of the participants felt that they learned more about the choices the government needed to make regarding the relaxation of lockdown measures through participating in the PVE, whereas around 20% disagreed with this proposition. Table 14 shows that awareness-raising about the consequences of relaxation options and the dilemmas the government faces was also cited by many participants as a strength of the method. Below, we list illustrative quotes of respondents that were positive about the awareness-raising ability of PVE:

*“This gives me a better understanding of the choice that politicians and policymakers face*.*”*

*“Everything was well-explained. The people that designed this research succeeded in showing that this is a choice with multiple dimensions instead of a simple choice. Great achievement that such a research is designed in such a small amount of time. It provides you as a participant with insights into the complexity of government choices*.*”*

*“I liked how you get insight into the consequences of relaxation options and the way that decisions on relaxing lockdown measures are interrelated*.*”*

*“This study increases the transparency of the trade-off that the government faces. Participants are also confronted with the consequences of their advices*.*”*

*“It made me think of how difficult these kinds of dilemmas are*.*”*

*“You experience the responsibility that people in government also experience”*.

Ideally, improved awareness improves the extent to which participants accept the final decision of the government and comply with government measures. To check this we asked respondents to evaluate the proposition:”Because the government involves me in this way, I am better able to accept the final decision of the government regarding the relaxation of lockdown measures between 20 May and 20 July.” 40% of the respondents agreed with this proposition and slightly more than 20% disagreed with it. Only a few respondents explicitly cited this as a strength of PVE. We also asked respondents on their opinion regarding the proposition: “since the government has asked for my advice, I will be more likely to adhere to the corona measures”. Our results show that only 18% thought that participating in a PVE would increase their compliance with lockdown measures.

Another potential benefit of PVE is that it provides an opportunity for participants to advise their government after experiencing a dilemma faced by policymakers. For reasons of limited space in the survey, we were not able to include a proposition which specifically asks participants how they perceived this specific characteristic of the PVE, but we asked them to respond to two propositions: “PVE is a good method for involving citizens in government decisions concerning the relaxation of government measures between 20 May and 20 July” and “The government should use this method more often for involving citizens in policymaking.” Around 80% of the respondents agreed with the proposition that PVE is a good method for involving citizens regarding this topic and 75% said that the government should use this method more often. Less-educated Dutch citizens are slightly more positive about the method than their highly educated counterparts.

Various participants cited some characteristics of PVE to explain why they thought it was a good method to transmit preferences of citizens to the government. Participants liked the fact that citizens were asked to provide advice based on insights regarding the consequences and that they were forced to make a choice between relaxation options. Moreover, participants liked that there was ample room to add nuances. Below, we provide illustrative quotes.

*“This setting allows participants to digest information about the consequences of government policies before they provide an advice. As a result, the outcomes are much more useful for government decision-makers than the preferences that people express on Facebook and Twitter*.*”*

*“You see the consequences of your advices. It is not a simple yes or no question without seeing the consequences like with the hopeless and useless idea of a referendum*.*”*

*“It is really good that people are asked to explain their choices because this ensures that people do not get away with pressing a few buttons based on their gut feeling*.*”*

*“The opportunity to provide written explanations. This allows you to express the nuances of your opinion that you cannot express with only making some choices between relaxation options*.*”*

Respondents also said that they liked that the PVE demonstrated that the government was open to the ideas of citizens.

*“I also like the fact that the government is open to the (good) ideas of its citizens. Thank you very much!”*

*“Nice way to involve people more directly in politics*.*”*

*“This allows people to communicate their concerns and worries. Now they use social media for this purpose, but I think it is very important and really useful to have a more formal place where people can blow off steam in a more productive way*.*”*

Only a handful of the 600 respondents mentioned as a strength that participating in the PVE gave them the feeling that their opinion counted. We also asked participants what weight politicians should assign to the outcomes of the PVE alongside the advice that politicians received from health experts. A minority of the participants (5%) thought that the advice given by citizens in the PVE should have a heavier weighting in the government’s decision-making than the advice given by experts. Conversely, 69% of participants opined that the expert advice should weigh heavier. The remaining 28% felt that the government should give both types of advice equal weighting. We think that it is interesting that citizens who participated in the PVE – who must have an above-average interest in participating in government decision-making – believe that more weight should be given to scientific advice than to the advice of citizens.

One potential downside of PVE is that the quality of preferences that people express is probably lower than preferences that people express after deliberation (such as is the case in mini-publics). It is, of course, difficult to directly verify the quality of the preferences of respondents, but as a surrogate, we asked respondents whether they were convinced of their advice. More than 70% of them responded positively to this proposition. Moreover, we asked respondents whether they changed their opinion due to participating in the PVE (about a third of the participants said that this was the case). In addition, respondents were asked to mention weaknesses of the method (or aspects that can be improved) and we only found one argument among the written answers of the 600 respondents we analysed which referred to limitations in terms of the ability to transmit preferences to the government via a PVE. A handful of participants criticised the fact that they could only make a distinction between different subsector (e.g. bars should be closed, but museums should be opened) in the written motivations and not in the primary choice tasks of the PVE. Other weaknesses were mentioned by a larger number of respondents: not possible to conduct the experiment via a smartphone, the profiles of relaxation options varied across respondents on their impact levels and pressure to the healthcare system (see section 4.2) and some respondents found this suspicious, some respondents found the survey too complex and, finally, respondents argued that the research team should bring the experiment under the attention by more people via advertisement to ensure that more people participate.

## 6 Conclusion and discussion

This paper reports about an attempt that was made in the Netherlands to involve about 30,000 Dutch citizens in policy decisions regarding relaxing lockdown measures between 20 May and 20 July 2020 through a Participatory Value Evaluation (PVE). Participants in the PVE were presented with eight possibilities for relaxing lockdown measures for this period, out of which they could make recommendations to the government. For each of these relaxation options, they received information regarding the option’s societal impact (e.g., increase in pressure on the health care system, an increase in deaths among people younger than 70 years and a decrease in the number of households with a long-term loss of income). The constraint that participants faced in the PVE was the maximum capacity of the healthcare system. They were not able to recommend a bundle of relaxation options that resulted in a greater than 50% increase in the pressure on the healthcare system. Subsequently, participants were asked which of the eight relaxation options should not be considered by the government. We carried out the PVE with two different samples. First, a random selection of 3,358 Dutch adults, who formed a representative sample of the Dutch population of 18 years and older. Second, we opened the PVE for the general public, which resulted in more than 26,000 participants within six days. The primary goal of this paper is to show what sorts of insights a PVE can provide to policymakers and other stakeholders who have to decide on COVID-19 policies. A secondary objective of this paper is to improve understanding towards the strengths and weaknesses of PVE in terms of involving citizens in crisis policymaking.

### 6.1 Main findings

Our results show that the majority of the participants in the PVE advised the government to relax lockdown measures, but not to the point at which the healthcare system becomes heavily overloaded. Participants in the ‘open PVE’ were inclined to support a somewhat more extensive relaxation of lockdown measures than the average Dutch citizen (participants in the representative PVE). From the choices respondents made in the PVE, we were able to infer the implicit trade-offs made by Dutch citizens between impacts of relaxation options. For instance, we find that a reduction of 100 deaths of persons below the age of 70 years and the reduction of 168 deaths of citizens older than 70 years are equally attractive. There is wide support among participants for re-opening contact professions and our results show that this option is popular in all segments of Dutch society. Conversely, we found little support for policy options that would relax restrictions for one specific group of citizens. The options “All restrictions lifted in Northern provinces” and “All restrictions lifted for people with immunity” can count on little support among the Dutch population at large. The low support for the option “All restrictions lifted in Northern provinces” is at odds with the message of a number of scientists who advocated this option in the weeks before we conducted the PVE^79^. Participants had a negative stance towards these relaxation options because they found it very important that the relaxation of lockdown measures leads to “unity” and not to “division”. They are afraid that the unity among Dutch people that currently exists – along with the support for corona-related government policies – will be lost if and when the Cabinet chooses to lift restrictions for a specific group of Dutch people. The importance of equal treatment is also identified in studies which examined Dutch citizens preferences regarding health policies before the outbreak of the coronavirus^79-80^. However, a clear contribution of our study is that Dutch citizens seem to think that it is unfair to distinguish policies between different regions, age groups and people who are (not) immune to COVID-19 – various respondents even labelled this as ‘discrimination’– whereas we did not identified any respondents who explicitly said that making distinctions between different sectors (contact professions, hospitality industry and other business) would be’unfair’. Another result that stands out is that 71% of the respondents who recommended the relaxation option “Nursing and care homes allow visitors” say that they will not experience any impacts from the implementation of this option. This suggests that involving large numbers of citizens in determining crisis policies might also increase empathy between individuals and foster an exchange of perspectives regarding ethical trade-offs^91^.

The choices made by participants in the PVE can be used as input for behaviourally-informed choice models which analyse people’s preferences for (the impacts of) relaxation policies. These preferences can, in turn, be used to rank options in terms of their desirability. We find that citizens consider a reduction of 100 deaths of persons below the age of 70 years to be equally attractive as a reduction of 168 deaths of citizens older than 70 years. We find that the optimal portfolio of relaxation policies consists of three strategies: re-open contact professions, re-open businesses (except the hospitality industry) and allow social contact between direct family members. An advantage of PVE is that sensitivity analyses can be conducted to explore how the desirability of policy options is affected by changes in impacts. These sensitivity analyses show that in a pessimistic scenario only re-opening contact professions is included in the optimal portfolio. In an optimistic scenario five out of eight relaxation policies are part of the optimal portfolio, excluding re-opening the hospitality industry, lifting restrictions for those with immunity and lifting restrictions for the Northern provinces.

In this paper, we listed various reasons why PVE could be an appealing participatory approach for involving citizens in policy decisions during a pandemic: 1) citizens can participate in a PVE online, which is appealing in times of social distancing; 2) a PVE can be deployed rapidly, which is important during a pandemic as governments have to respond quickly to new developments; 3) the design of a PVE can adopt other constraints than only public budget; 4) PVE provides information to policymakers about the extent to which the desirability of policy options is affected by the impacts of the policy options; 5) PVE allows citizens to transmit new ideas, arguments, values and conditions to decision-makers; 6) PVE enables citizens to participate in multi-dimensional policy issues that do not lend themselves to a simple ‘yes’ or ‘no’ or the allocation of a constrained amount of scarce public resources; 7) PVE lets citizens experience intricate government dilemmas, increasing their understanding of the impacts of proposed measures and potentially increasing levels of acceptance and compliance.

In this paper, we establish that the first five potential benefits of the method were realised in this PVE. Citizens could participate online and the PVE was deployed rapidly (designing process started 9 April, 2020 and results were shared with policymakers 6 May, 2020). The PVE adopted another constraint than the public budget (maximum pressure on the health care system), we showed that the PVE provided information to policymakers about the extent to which the desirability of policy options is affected by the impacts of the policy options and the PVE allowed citizens to transmit new ideas, arguments, values and conditions to policymakers. Policymakers can embed these new ideas, arguments, values and conditions in their policies, and the quantitative results produced by the PVE – such as the ranking of relaxation options – can inform their prioritisations. Moreover, the outcomes of the PVE provides policymakers with information about the effectiveness of existing policies. For instance, many respondents said that they themselves (or other people) were already violating the rule that family members from another household cannot have social contact.

We think that we can safely conclude that we partially realised the sixth and seventh appealing characteristics of PVE. Almost 60% of respondents said that they became more aware of the consequences of relaxation options and the dilemmas the government faces (instrumental rationale for participation). Almost 80% of participants stated that PVE is a good method to let citizens participate in government decision-making on lifting lockdown measures. Participants liked the fact that they were asked to provide advice while evaluating relaxation options in relation to each other and being informed about the consequences of the options. Participants also appreciated that they were forced to make a choice between relaxation options and that there was ample room to add nuances. That said, our results do not show convincingly that respondents would also comply to a higher level with public health measures simply because they participated in our study (only 18% said that this was the case) or that participation in the PVE would increase their acceptance of the lockdown policies of the government (only 40% argued that participation in the PVE would increase their acceptance).

### 6.2 Limitations and further research

One major benefit of a PVE is that it can be deployed rapidly, but at the same time many limitations of our study were caused by the short timeframe. The study was designed in 20 days and the data was collected and analysed in 7 days. It goes without saying that the quality of our study would have been higher if we had had more time to design the study and analyse the data. Had this been the case, we probably would have analysed the written motivations of a larger number of respondents to provide policymakers with an even larger set of new ideas, conditions and values that the respondents aimed to transmit to their government. The fact that we were not able to analyse all the written motivations is problematic because participants cited the fact that PVE provides a lot of opportunities to explain their advises and to add nuances as a key strength of PVE. We believe that this shortcoming can be alleviated by analysing the qualitative data faster and more systematically through natural language processing and using a larger group of annotators. Another limitation of our study that was caused by time pressure is that we were not able to finalise a mobile version in time. Moreover, on the first day of our data collection, the PVE went offline due to lack of server capacity. With a mobile version and enough server capacity in place, we believe that the number of participants would have been substantially higher. Despite these limitations, we believe that the PVE can serve as an example for policymakers and academics of what can realistically be achieved in terms of involving the public in crisis policymaking^12^. PVE is probably a cheaper and more efficient alternative to live experimentation – that is, imposing policies on citizens and seeing what sticks^12^. Another limitation of our study concerns its generalisation to other contexts. This research is only a temporary glance into Dutch citizens’ preferences concerning the relaxation of lockdown measures in late April 2020. Citizens in different countries and cultures might have different preferences. Furthermore, preferences can shift as the severity of the pandemic, individual experiences and risk perceptions and the efficacy of pharmaceutical and non-pharmaceutical measures evolve over time. It would be interesting to repeat the PVE in different contexts (time and location) to explore its general is ability in terms of outcomes and the way that the method is perceived by participants.

Even though there is no point of comparison, we reason that the quality of preferences that people express in the PVE is probably lower than preferences that they express after deliberation (such as is the case in mini-publics). This is because citizens’ interests, preferences and perceptions of a crisis situation are not fixed but subject to discursive challenges. In the PVE, respondents were provided with information on the policy alternatives that were on the government’s table, but – as far as we know – most of them studied this information individually, without the opportunity to ask questions of experts, discuss implications with other groups of people, and so forth^66^. Not only is preference formation an inherently social and dynamic process, so is the adherence to social distancing recommendations during the COVID-19 pandemic^92^. Therefore, as mentioned earlier, various scholars argue that deliberation with others is decisive for preference formation^66-67^. When citizens deliberate, they can expand their knowledge, including both their own self-understanding and their collective understanding of what will best serve other affected groups^93^. Moreover, empirical studies show that individuals interacting with one another generally outperform groups of unconnected individuals^94^. Hence, enriching PVE experiments with deliberative elements (e.g., group discussion, consulting expert witnesses or a forum) may contribute to well-formed preferences in the case of unfamiliar and complex government policies and may even increase adherence to subsequent government measures^66-67^. Augmenting PVE with deliberative elements will allow participating citizens to learn from each other, to form reasoned opinions and to evaluate positions, thereby ironing out critiques of the individual approach to preference formation. It is important to investigate the extent to which the beneficial aspects of social interaction outweigh potential downsides such as social bias, herding and groupthink to ensure that social interaction leads to the ‘wisdom of the crowd’ instead of the ‘madness of the mob’^95^. For the same reason, we believe that PVE is merely one of several ways to involve citizens in crisis policymaking, and might complement other public participation methods. In our view, PVE could be optimally used jointly with deliberative methods, such as mini-publics. For instance, a mini-public could be used in the design stage of the PVE (selecting relaxation options which are included in the PVE) and could then also be asked to translate the results of the PVE into policy recommendations.

A final promising avenue for further research would be to study how the results of the PVE could (better) fit in political decision-making processes. In the context of this PVE, we had contact with civil servants, but we were not in contact with the Dutch parliament and we aren’teven aware of whether they received our report. It would be interesting to study how a PVE should be institutionalized in a representative democracy, also considering the fact that only 5% of participants in the PVE itself demanded that their advice as citizens should count for more than that of experts.

## Data Availability

Data will become available after publication of the paper

## Acknowledgements

We are grateful to Kantar Public for charging us less than their usual rate per obtained respondent. We received valuable input from the following individuals during the design of the PVE, the implementation of the PVE and the analysis of the data: Marion Collewet, Sjoerd Jenninga, Selma van Delft, Julia Kooijman, Lionel Kaptein, Lisa Volberda, Enrico Liscio, Pradeep Murukannaiah, Perry Borst, Shannon Spruit, Ardine de Wit, Adrienne Rotteveel, Mattijs Lambooij, Anita Suijkerbuijk, Paul van Gils, Toep van Dijk, Tomas Peeters, Suzanne Pietersma, Denny Borsboom, Tessa Blanken, Job van Exel, Sake de Vlas, Marcel Jansen, Vincent van Petten, Nienke van der Haak, Hans Heesterbeek, and Rob Kooij.

## Appendix 1: Experimental design process.

The first part of the experimental design consisted of defining the possible impact levels and pressure on the healthcare system of each relaxation option, based on the feedback and information that was obtained in the PVE design process. Table A1.1 summarizes each possible impact including the increase in the pressure on the health care system caused by the relaxation option.

Ideally, an experimental design should consider all possible combinations of impact and pressure levels, in order to capture information from all the possible profiles of relaxation strategies from participants choices in the PVE. This is called in literature as a “full-factorial design”. However, collecting data for such design is often non-tractable, because the number of combinations explodes even for small numbers of relaxation strategies, impacts, and impact/pressure levels. For this PVE, a full factorial design is composed of more than 1.59 ∗ 10^26^ combinations.

**Table A1:**
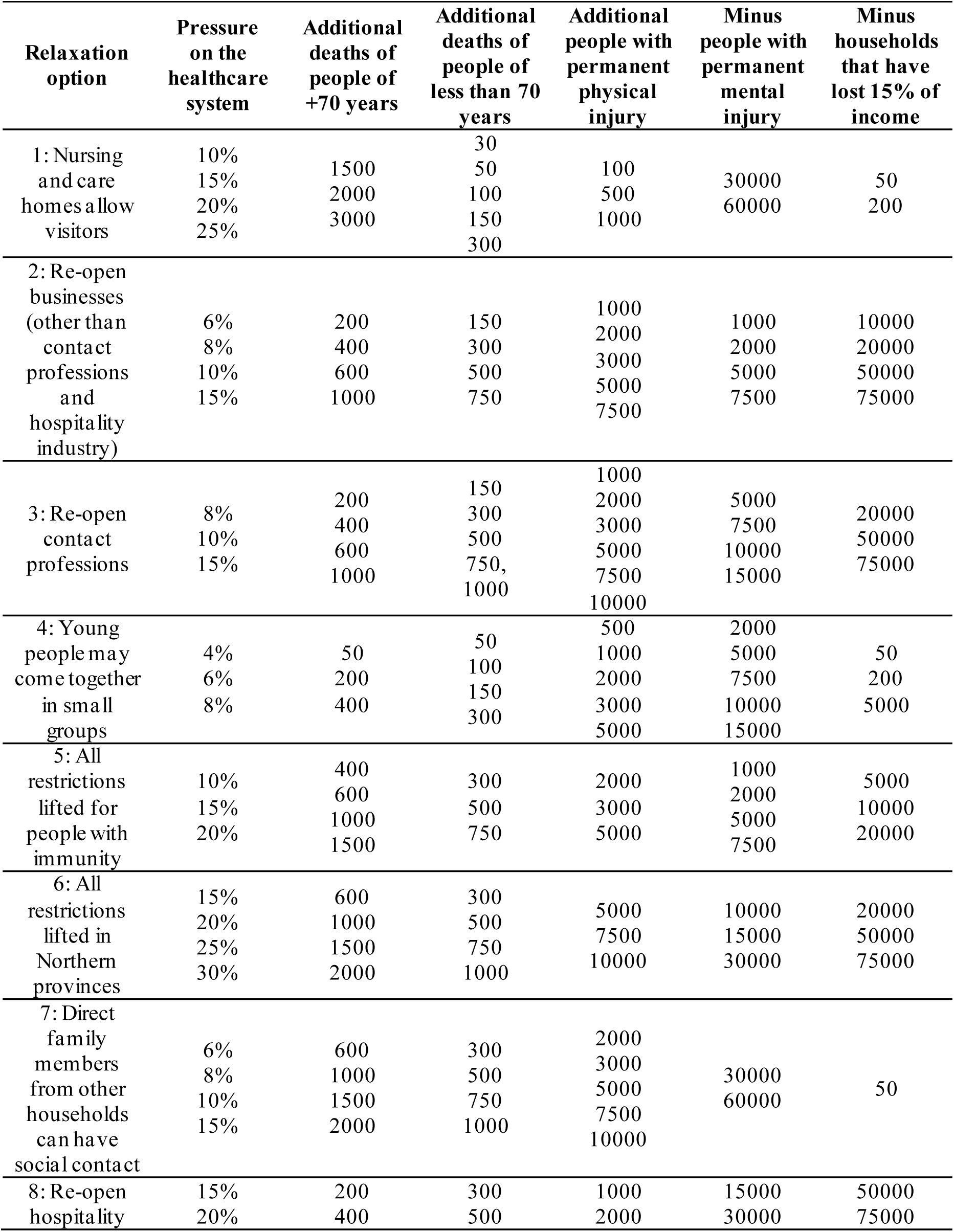
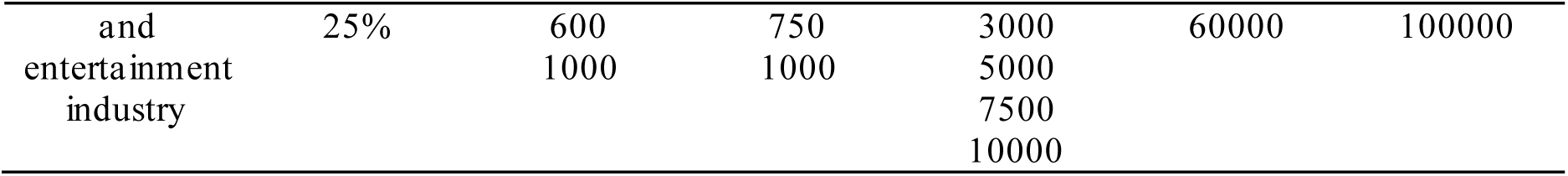
Possible impact levels and pressure on the healthcare system for each relaxation option.

There are several solutions to reduce the number of combinations of the experimental design. The first and most intuitive one is to take a random number of profiles (defined by the researcher) of the full factorial design. This is called in literature as a “fractional factorial design”. A problem of this approach is that it artificially increases the correlation level between impact/pressure levels of the experimental design. In turn, this increased correlation has an impact on the possibility of extracting information related to preferences for impacts (i.e. taste parameters) using econometric models. In light of this, the construction of a reduced experimental design should ensure that the correlation between attributes is minimal.

The experimental design of this PVE aims to obtain a tractable number of profiles, at the same time that the correlation between impact/pressure levels is minimized. In particular, we aim to minimize the maximum value of the correlation matrix between impact/pressure levels of the design:

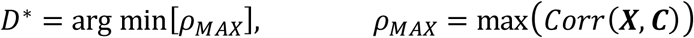

Where *D*^∗^ is the optimal design matrix, ***X*** is a *N* × *J* × *K* matrix of impact levels for *N* profiles, *J* relaxation options, and *K* impacts; ***C*** is a *N* × *J* matrix of pressure levels for each profile and relaxation option. We call this type of designs as “min-max correlation” designs.

We developed an algorithm that creates min-max correlation designs by iteratively selecting impact/pressure levels, evaluating on each step whether the max-correlation is reduced. This algorithm is described as follows:

- <u>Step 0 (Definition of inputs):</u> Define the set of possible impact and pressure levels for each relaxation option. Define *N* as the number of required profiles.
- <u>Step 1 (Definition of initial candidate design):</u> Construct 10 designs with N profiles each one, by taking random levels of impact and pressure levels from the set defined on Step 0. Keep the design with the smallest max-correlation value and call it as “(initial) candidate design”.
- <u>Step 2 (Replacement):</u> Set a random impact/pressure from the candidate design, and replace its value with a random impact/pressure value from the set defined in Step 0, and call it as the new “candidate design”.
- <u>Step 3 (Evaluation):</u> Compute the max-correlation value of the candidate design.
- <u>Step 4 (Decision):</u> If the max-correlation value of the new candidate design is reduced, the change is kept and go back to Step 2. Otherwise, the change is reverted and go back to Step 2.

This algorithm is conducted until a certain number of iterations without improvement is reached, or until a certain amount of time. Then, the last stored design is called as the optimal design. For this PVE, we ran the algorithm for 10 minutes, although we observed no further improvement after 3 minutes approximately. Finally, we introduced additional constraints to the replacement step in the algorithm in order to avoid that a possible impact/pressure level does not appear in the optimal design.

## Appendix 2: Descriptive information for sociodemographic variables used in Figure 1 and Table 5

**Table A2:**
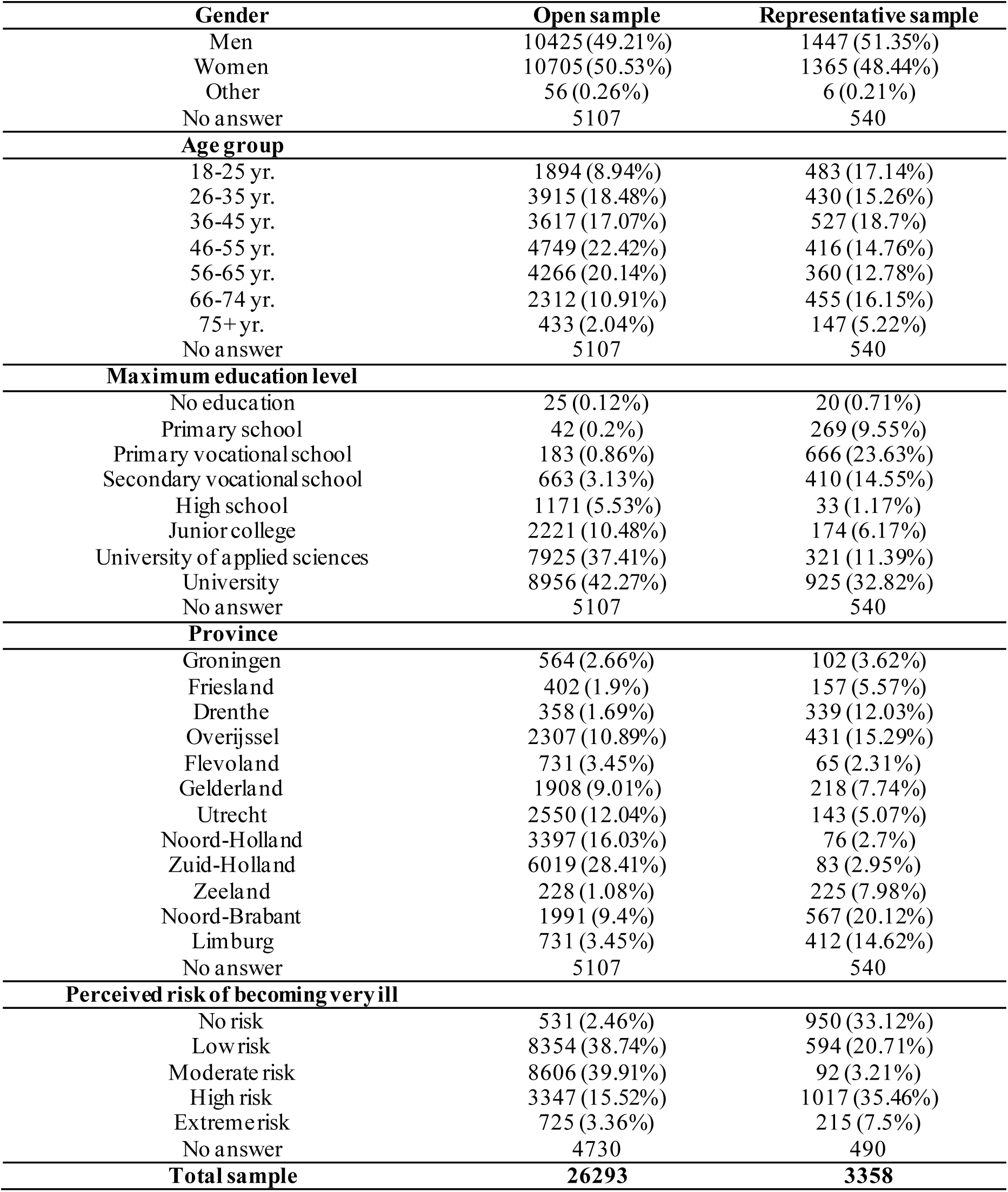
Frequency of sociodemographic variables used in Figure 1 and Table 5.

## Appendix 3: MDCEV estimates and optimal portfolio for separate samples

**Table A3.1:** MDCEV model estimates. Separate samples.

|  | Open sample | Representative sample |
| --- | --- | --- |
| **Policy-specific constants:** |  |  |
| 1: Nursing and care homes allow visitors | 2.6865***<br>(0.0297) | 2.7219***<br>(0.0764) |
| 2: Re-open businesses (other than contact professions and hospitality industry) | 2.6451***<br>(0.0233) | 2.4132***<br>(0.0556) |
| 3: Re-open contact professions | 3.2382***<br>(0.0276) | 2.8500***<br>(0.0631) |
| 4: Young people may come together in small groups | 1.8825***<br>(0.0142) | 1.6317***<br>(0.0357) |
| 5: All restrictions lifted for people with immunity | 1.5608***<br>(0.0211) | 1.9986***<br>(0.0533) |
| 6: All restrictions lifted in Northern provinces | 1.5954***<br>(0.0342) | 2.0641***<br>(0.0809) |
| 7: Direct family members from other households can have social contact | 2.4893***<br>(0.0294) | 2.6784***<br>(0.0748) |
| 8: Re-open hospitality and entertainment industry | 2.7346***<br>(0.0376) | 2.4078***<br>(0.0857) |
| **Taste parameters:** |  |  |
| Additional 10.000 deaths of people of +70 years | -0.4123***<br>(0.0945) | -1.1009***<br>(0.2308) |
| Additional 10.000 deaths of people of less than 70 years | -0.9295***<br>(0.1933) | -0.4503<br>(0.4380) |
| Additional 10.000 people with permanent physical injury | -0.1033***<br>(0.0174) | -0.1481***<br>(0.0434) |
| Minus 10.000 people with permanent mental injury | 0.0023<br>(0.0037) | -0.0121<br>(0.0094) |
| Minus 10.000 households that have lost 15% of income | 0.0094***<br>(0.0026) | -0.0042<br>(0.0060) |
| Observations | 26293 | 3358 |
| Log-likelihood | -127928.8123 | -16499.3413 |
| AIC | 255831.6246 | 32972.6826 |
| BIC | 255725.3229 | 32893.1343 |

**Note:** Standard errors in parenthesis. **Statistical significance:** \*\*\* p < 0.001, \*\* p < 0.01, \* p < 0.05

**Table A3.2:** Optimal portfolios of relaxation options. Separate samples.

|  | Open sample |  |  | Representative sample |  |  |
| --- | --- | --- | --- | --- | --- | --- |
|  | Avg. | Pessim. | Optim | Avg. | Pessim. | Optim |
| 1: Nursing and care homes allow visitors |  |  | X |  |  | X |
| 2: Re-open businesses (other than contact professions and hospitality industry) | X |  | X |  |  | X |
| 3: Re-open contact professions | X | X | X | X |  | X |
| 4: Young people may come together in small groups |  |  | X |  |  | X |
| 5: All restrictions lifted for people with immunity |  |  |  |  |  |  |
| 6: All restrictions lifted in Northern provinces |  |  |  |  |  |  |
| 7: Direct family members from other households can have social contact | X |  | X | X |  | X |
| 8: Re-open hospitality and entertainment industry |  |  | X |  |  |  |
| **Pressure to the healthcare system** | **31.6%** | **15%** | **49%** | **21.8%** | **0%** | **34%** |

## Appendix 4: Quantitative results and impact/pressure levels used for sensitivity analysis. Sample of provinces of Friesland, Groningen and Drenthe (the Northern provinces)

**Table A4.1:** MDCEV model estimates. Sample for individuals who live in the Northern provinces of Friesland, Groningen and Drenthe.

|  | Estimates |
| --- | --- |
| **Baseline utility of relaxation strategies:** |  |
| 1: Nursing and care homes allow visitors | 2.7526***<br>(0.1085) |
| 2: Re-open businesses (other than contact professions and hospitality industry) | 2.3516***<br>(0.0797) |
| 3: Re-open contact professions | 2.9320***<br>(0.0923) |
| 4: Young people may come together in small groups | 1.7161***<br>(0.0511) |
| 5: All restrictions lifted for people with immunity | 1.4744***<br>(0.0803) |
| 6: All restrictions lifted in Northern provinces | 2.2522***<br>(0.1139) |
| 7: Direct family members from other households can have social contact | 2.3574***<br>(0.1072) |
| 8: Re-open hospitality and entertainment industry | 2.4172***<br>(0.1219) |
| **Impact effects:** |  |
| Additional 10.000 deaths of people of +70 years | -0.7926*<br>(0.3298) |
| Additional 10.000 deaths of people of less than 70 years | -0.7957<br>(0.6428) |
| Additional 10.000 people with permanent physical injury | -0.0492<br>(0.0616) |
| Minus 10.000 people with permanent mental injury | 0.0042<br>(0.0132) |
| Minus 10.000 households that have lost 15% of income | 0.0007<br>(0.0085) |
| Observations | 1645 |
| Log-likelihood | -8073.5348 |
| AIC | 16121.0695 |
| BIC | 16050.7981 |

**Table A4.2:** Optimal portfolios of relaxation options. Sample for individuals who live in the Northern provinces of Friesland, Groningen and Drenthe.

|  | Averages | Pessimistic | Optimistic |
| --- | --- | --- | --- |
| 1: Nursing and carehomes allow visitors |  |  | X |
| 2: Re-open businesses (other than contact professions and hospitality industry) |  |  | X |
| 3: Re-open contact professions | X | X | X |
| 4: Young people may come together in small groups |  |  | X |
| 5: All restrictions lifted for people with immunity |  |  |  |
| 6: All restrictions lifted in Northern provinces |  |  |  |
| 7: Direct family members from other households can have social contact |  |  | X |
| 8: Re-open hospitality and entertainment industry |  |  |  |
| **Pressure to the healthcare system** | **11.4%** | **15%** | **34%** |

**Table A4.3:** Impact levels used for optimal portfolio computation for three scenarios. Sample for individuals who live in the Northern provinces of Friesland, Groningen and Drenthe.

| Impact | Relaxation strategy | Average | Conservative | Optimistic |
| --- | --- | --- | --- | --- |
| Additional deaths of people of +70 years | Nursing and care homes allow visitors | 2185.41 | 3000 | 1500 |
|  | Re-open businesses (other than contact professions and hospitality industry) | 712.58 | 1000 | 200 |
|  | Re-open contact professions | 591.85 | 1000 | 200 |
|  | Young people may come together in small groups | 251.52 | 400 | 50 |
|  | All restrictions lifted for people with immunity | 1060.49 | 1500 | 400 |
|  | All restrictions lifted in Northern provinces | 1218.18 | 2000 | 600 |
|  | Direct family members from other households can have social contact | 1127.54 | 2000 | 600 |
|  | Re-open hospitality and entertainment industry | 567.29 | 1000 | 200 |
| Additional deaths of people of less than 70 years | Nursing and care homes allow visitors | 130.70 | 300 | 30 |
|  | Re-open businesses (other than contact professions and hospitality industry) | 468.33 | 750 | 150 |
|  | Re-open contact professions | 576.66 | 1000 | 150 |
|  | Young people may come together in small groups | 161.25 | 300 | 50 |
|  | All restrictions lifted for people with immunity | 554.89 | 750 | 300 |
|  | All restrictions lifted in Northern provinces | 641.82 | 1000 | 300 |
|  | Direct family members from other households can have social contact | 605.74 | 1000 | 300 |
|  | Re-open hospitality and entertainment industry | 570.67 | 1000 | 300 |
| Additional people with permanent physical injury | Nursing and care homes allow visitors | 619.88 | 1000 | 100 |
|  | Re-open businesses (other than contact professions and hospitality industry) | 4049.24 | 7500 | 1000 |
|  | Re-open contact professions | 4612.16 | 10000 | 1000 |
|  | Young people may come together in small groups | 2636.17 | 5000 | 500 |
|  | All restrictions lifted for people with immunity | 3430.40 | 5000 | 2000 |
|  | All restrictions lifted in Northern provinces | 7500 | 10000 | 5000 |
|  | Direct family members from other households can have social contact | 5422.80 | 10000 | 2000 |
|  | Re-open hospitality and entertainment industry | 4859.27 | 10000 | 1000 |
| Reduction of people with permanent mental injury | Nursing and care homes allow visitors | 41069.91 | 30000 | 60000 |
|  | Re-open businesses (other than contact professions and hospitality industry) | 4031 | 1000 | 7500 |
|  | Re-open contact professions | 9575.99 | 5000 | 15000 |
|  | Young people may come together in small groups | 6765.96 | 2000 | 10000 |
|  | All restrictions lifted for people with immunity | 3676.60 | 1000 | 7500 |
|  | All restrictions lifted in Northern provinces | 17054.71 | 10000 | 30000 |
|  | Direct family members from other households can have social contact | 46778.12 | 30000 | 60000 |
|  | Re-open hospitality and entertainment industry | 42246.20 | 15000 | 60000 |
| Reduction of households that have lost 15% of income | Nursing and care homes allow visitors | 141.19 | 50 | 200 |
|  | Re-open businesses (other than contact professions and hospitality industry) | 38270.52 | 10000 | 75000 |
|  | Re-open contact professions | 50434.65 | 20000 | 75000 |
|  | Young people may come together in small groups | 1371.37 | 50 | 5000 |
|  | All restrictions lifted for people with immunity | 12167.17 | 5000 | 20000 |
|  | All restrictions lifted in Northern provinces | 50206.69 | 20000 | 75000 |
|  | Direct family members from other households can have social contact | 50 | 50 | 50 |
|  | Re-open hospitality and entertainment industry | 75516.72 | 50000 | 100000 |

**Table A4.4:** Pressure to the healthcare system used for optimal portfolio computation for three scenarios. Sample for individuals who live in the Northern provinces of Friesland, Groningen and Drenthe.

| Relaxation strategy | Average | Conservative | Optimistic |
| --- | --- | --- | --- |
| Nursing and care homes allow visitors | 18.02 | 25 | 10 |
| Re-open businesses (other than contact professions and hospitality industry) | 9.87 | 15 | 6 |
| Re-open contact professions | 11.49 | 15 | 8 |
| Young people may come together in small groups | 6.59 | 8 | 4 |
| All restrictions lifted for people with immunity | 15.19 | 20 | 10 |
| All restrictions lifted in Northern provinces | 22.23 | 30 | 15 |
| Direct family members from other households can have social contact | 10.11 | 15 | 6 |
| Re-open hospitality and entertainment industry | 19.09 | 25 | 15 |

## Appendix 5: Impact/pressure levels used for sensitivity analysis, for each type of sample.

**Table A5.1:** Impact levels used for optimal portfolio computation for three scenarios. Open sample.

| Impact | Relaxation strategy | Average | Conservative | Optimistic |
| --- | --- | --- | --- | --- |
| Additional deaths of people of +70 years | Nursing and care homes allow visitors | 2215.27 | 3000 | 1500 |
|  | Re-open businesses (other than contact professions and hospitality industry) | 714.07 | 1000 | 200 |
|  | Re-open contact professions | 593.74 | 1000 | 200 |
|  | Young people may come together in small groups | 248.61 | 400 | 50 |
|  | All restrictions lifted for people with immunity | 1063.01 | 1500 | 400 |
|  | All restrictions lifted in Northern provinces | 1220.99 | 2000 | 600 |
|  | Direct family members from other households can have social contact | 1120.51 | 2000 | 600 |
|  | Re-open hospitality and entertainment industry | 569.73 | 1000 | 200 |
| Additional deaths of people of less than 70 years | Nursing and care homes allow visitors | 129.62 | 300 | 30 |
|  | Re-open businesses (other than contact professions and hospitality industry) | 471.64 | 750 | 150 |
|  | Re-open contact professions | 573.65 | 1000 | 150 |
|  | Young people may come together in small groups | 161.98 | 300 | 50 |
|  | All restrictions lifted for people with immunity | 548.56 | 750 | 300 |
|  | All restrictions lifted in Northern provinces | 638.18 | 1000 | 300 |
|  | Direct family members from other households can have social contact | 597.84 | 1000 | 300 |
|  | Re-open hospitality and entertainment industry | 569.27 | 1000 | 300 |
| Additional people with permanent physical injury | Nursing and care homes allow visitors | 614.30 | 1000 | 100 |
|  | Re-open businesses (other than contact professions and hospitality industry) | 3961.32 | 7500 | 1000 |
|  | Re-open contact professions | 4611.11 | 10000 | 1000 |
|  | Young people may come together in small groups | 2592.76 | 5000 | 500 |
|  | All restrictions lifted for people with immunity | 3404.78 | 5000 | 2000 |
|  | All restrictions lifted in Northern provinces | 7522.15 | 10000 | 5000 |
|  | Direct family members from other households can have social contact | 5540.20 | 10000 | 2000 |
|  | Re-open hospitality and entertainment industry | 4945.23 | 10000 | 1000 |
| Reduction of people with permanent mental injury | Nursing and care homes allow visitors | 40728.71 | 30000 | 60000 |
|  | Re-open businesses (other than contact professions and hospitality industry) | 4059.84 | 1000 | 7500 |
|  | Re-open contact professions | 9532.19 | 5000 | 15000 |
|  | Young people may come together in small groups | 6756.25 | 2000 | 10000 |
|  | All restrictions lifted for people with immunity | 3721.41 | 1000 | 7500 |
|  | All restrictions lifted in Northern provinces | 17249.46 | 10000 | 30000 |
|  | Direct family members from other households can have social contact | 46466.74 | 30000 | 60000 |
|  | Re-open hospitality and entertainment industry | 42606.21 | 15000 | 60000 |
| Reduction of households that have lost 15% of income | Nursing and care homes allow visitors | 140.75 | 50 | 200 |
|  | Re-open businesses (other than contact professions and hospitality industry) | 39210.82 | 10000 | 75000 |
|  | Re-open contact professions | 49987.83 | 20000 | 75000 |
|  | Young people may come together in small groups | 1408.73 | 50 | 5000 |
|  | All restrictions lifted for people with immunity | 12051.50 | 5000 | 20000 |
|  | All restrictions lifted in Northern provinces | 49546.08 | 20000 | 75000 |
|  | Direct family members from other households can have social contact | 50 | 50 | 50 |
|  | Re-open hospitality and entertainment industry | 75771.12 | 50000 | 100000 |

**Table A5.2:**
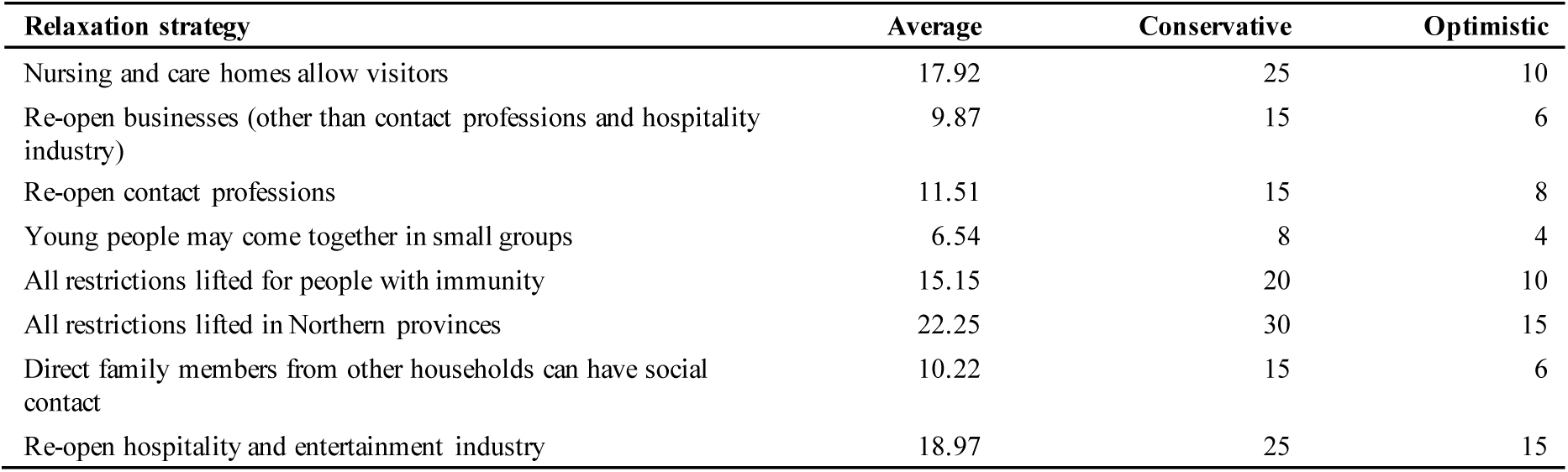
Pressure to the healthcare system used for optimal portfolio computation for three scenarios. Open sample.

**Table A5.3:** Impact levels used for optimal portfolio computation for three scenarios. Representative sample.

| Impact | Relaxation strategy | Average | Conservative | Optimistic |
| --- | --- | --- | --- | --- |
| Additional deaths of people of +70 years | Nursing and care homes allow visitors | 2223.65 | 3000 | 1500 |
|  | Re-open businesses (other than contact professions and hospitality industry) | 706.37 | 1000 | 200 |
|  | Re-open contact professions | 588.92 | 1000 | 200 |
|  | Young people may come together in small groups | 250.07 | 400 | 50 |
|  | All restrictions lifted for people with immunity | 1062.63 | 1500 | 400 |
|  | All restrictions lifted in Northern provinces | 1221.11 | 2000 | 600 |
|  | Direct family members from other households can have social contact | 1110.75 | 2000 | 600 |
|  | Re-open hospitality and entertainment industry | 570.40 | 1000 | 200 |
| Additional deaths of people of less than 70 years | Nursing and care homes allow visitors | 129.01 | 300 | 30 |
|  | Re-open businesses (other than contact professions and hospitality industry) | 471.86 | 750 | 150 |
|  | Re-open contact professions | 569.57 | 1000 | 150 |
|  | Young people may come together in small groups | 161.49 | 300 | 50 |
|  | All restrictions lifted for people with immunity | 549.24 | 750 | 300 |
|  | All restrictions lifted in Northern provinces | 639.90 | 1000 | 300 |
|  | Direct family members from other households can have social contact | 597.29 | 1000 | 300 |
|  | Re-open hospitality and entertainment industry | 569.88 | 1000 | 300 |
| Additional people with permanent physical injury | Nursing and care homes allow visitors | 604.05 | 1000 | 100 |
|  | Re-open businesses (other than contact professions and hospitality industry) | 4010.42 | 7500 | 1000 |
|  | Re-open contact professions | 4625.67 | 10000 | 1000 |
|  | Young people may come together in small groups | 2580.85 | 5000 | 500 |
|  | All restrictions lifted for people with immunity | 3410.66 | 5000 | 2000 |
|  | All restrictions lifted in Northern provinces | 7510.42 | 10000 | 5000 |
|  | Direct family members from other households can have social contact | 5498.66 | 10000 | 2000 |
|  | Re-open hospitality and entertainment industry | 4864.35 | 10000 | 1000 |
| Reduction of people with permanent mental injury | Nursing and care homes allow visitors | 40640.26 | 30000 | 60000 |
|  | Re-open businesses (other than contact professions and hospitality industry) | 4032.61 | 1000 | 7500 |
|  | Re-open contact professions | 9586.81 | 5000 | 15000 |
|  | Young people may come together in small groups | 6745.09 | 2000 | 10000 |
|  | All restrictions lifted for people with immunity | 3725.58 | 1000 | 7500 |
|  | All restrictions lifted in Northern provinces | 17374.93 | 10000 | 30000 |
|  | Direct family members from other households can have social contact | 46340.08 | 30000 | 60000 |
|  | Re-open hospitality and entertainment industry | 42480.64 | 15000 | 60000 |
| Reduction of households that have lost 15% of income | Nursing and care homes allow visitors | 140.54 | 50 | 200 |
|  | Re-open businesses (other than contact professions and hospitality industry) | 38941.33 | 10000 | 75000 |
|  | Re-open contact professions | 50064.03 | 20000 | 75000 |
|  | Young people may come together in small groups | 1466.24 | 50 | 5000 |
|  | All restrictions lifted for people with immunity | 12182.85 | 5000 | 20000 |
|  | All restrictions lifted in Northern provinces | 50023.82 | 20000 | 75000 |
|  | Direct family members from other households can have social contact | 50 | 50 | 50 |
|  | Re-open hospitality and entertainment industry | 75707.27 | 50000 | 100000 |

**Table A5.4:** Pressure to the healthcare system used for optimal portfolio computation for three scenarios. Representative sample.

| Relaxation strategy | Average | Conservative | Optimistic |
| --- | --- | --- | --- |
| Nursing and care homes allow visitors | 17.99 | 25 | 10 |
| Re-open businesses (other than contact professions and hospitality industry) | 9.91 | 15 | 6 |
| Re-open contact professions | 11.49 | 15 | 8 |
| Young people may come together in small groups | 6.52 | 8 | 4 |
| All restrictions lifted for people with immunity | 15.17 | 20 | 10 |
| All restrictions lifted in Northern provinces | 22.21 | 30 | 15 |
| Direct family members from other households can have social contact | 10.30 | 15 | 6 |
| Re-open hospitality and entertainment industry | 19.02 | 25 | 15 |

## References

1. Acemoglu D, Robinson JA. Why nations fail: The origins of power, prosperity, and poverty. Currency; 2012.

2. Weible CM, Nohrstedt D, Cairney P, Carter DP, Crow DA, Durnová AP, et al. COVID-19 and the policy sciences: initial reactions and perspectives. Policy Sci. 2020;53(2):1–17.

3. Vaughan E, Tinker T. Effective health risk communication about pandemic influenza for vulnerable populations. Am J Public Health. 2009;99 Suppl 2(S2):S324 –32

4. OECD. The territorial impact of COVID-19: Managing the crisis across levels of government [Internet]. Oecd.org. [cited 2020 Nov 3]. Available from: https://www.oecd.org/coronavirus/policy-responses/the-territorial-impact-of-COVID-19-managing-the-crisis-across-levels-of-government-d3e314e1/

5. Lavazza A, Farina M. The role of experts in the Covid-19 pandemic and the limits of their epistemic authority in democracy. Front Public Health. 2020;8:356.

6. Cairney P. The politics of evidence based policymaking. London: Palgrave; 2016

7. Dostal JM. Governing under pressure: German policy making during the Coronavirus crisis. Polit Q. 2020;91(3):542–52.

8. Sabat I, Neuman-Böhme S, Varghese NE, Barros PP, Brouwer W, van Exel J, et al. United but divided: Policy responses and people’s perceptions in the EU during the COVID-19 outbreak. Health Policy [Internet]. 2020; Available from: http://dx.doi.org/10.1016/j.healthpol.2020.06.009

9. Dryzek JS, Bächtiger A, Chambers S, Cohen J, Druckman JN, Felicetti A, et al. The crisis of democracy and the science of deliberation. Science. 2019;363(6432):1144 –6.

10. Ryan M, Smith G. Defining mini-publics. Deliberative mini-publics: Involving citizens in the democratic process, 9–26. In: Grönlund K, Bächtiger A, Setälä M, editors. Ecpr Press; 2014.

11. Open Government Partnership (OGP). Collecting open Government Approaches to COVID-19 [Internet]. Opengovpartnership.org. 2020 [cited 2020 Nov 3]. Available from: https://www.opengovpartnership.org/collecting-open-government-approaches-to-COVID-19.

12. Pearse H. Deliberation, citizen science and covid-19. Polit Q. 2020;91(3):571 –7.

13. Bernier H. Lessons learned from implementinga multi-year, multi-projectpublic engagement initiative to betterinform governmental public health policy decisions. J Participat Med May. 2014;22:68.

14. Schoch-Spana M, Franco C, Nuzzo JB, Usenza C, Working Group on Community Engagement in Health Emergency Planning. Community engagement: leadership tool for catastrophic health events. Biosecur Bioterror. 2007;5(1):8–25.

15. Schoch-Spana M, Chamberlain A, Franco C, Gross J, Lam C, Mulcahy A, et al. Disease, disaster, and democracy: the public’s stake in health emergency planning. Biosecur Bioterror. 2006;4(3):313–9.

16. Farrell H. The political economy of trust: Institutions, interests, and inter-firm cooperation in Italy and Germany. Cambridge University Press; 2009.

17. Lewis-Kraus G. How to Make Government Trustworthy Again. Why have some Asian countries controlled their outbreaks so well? It’s because authorities have earned their citizens’ confidence. Retrieved at: https://www.wired.com/story/how-to-make-government-trustworthy-again/. Wired [Internet]. 2020 Jun 18 [cited 2020 Nov 3]; Available from: https://www.wired.com/story/how-to-make-government-trustworthy-again/.

18. Fung A, Wright O. Thinking about Empowered Participatory Governance. Deepening democracy: Institutional innovations in empowered participatory governance. 2003;4(3).

19. Lee S, Hwang C, Moon MJ. Policy learning and crisis policy-making: quadruple-loop learning and COVID-19 responses in South Korea. Policy Soc. 2020;39(3):363–81.

20. Delgado A, Lein Kjølberg K, Wickson F. Public engagement coming of age: From theory to practice in STS encounters with nanotechnology. Public Underst Sci. 2011;20(6):826– 45.

21. Kingsley P. Serbia protests meet violent response in Europe’s 1st major virus unrest. The New York times [Internet]. 2020 Jul 9 [cited 2020 Nov 3]; Available from: https://www.nytimes.com/2020/07/08/world/europe/serbia-protests-coronavirus.html

22. Povoledo E, Minder R, Kwai I. Protesters in Italy and Spain clash with police as they call for ‘freedom’ from virus restrictions. The New York times [Internet]. 2020 Oct 27 [cited 2020 Nov 3]; Available from: https://www.nytimes.com/2020/10/27/world/protesters-in-italy-and-spain-clash-with-police-as-they-call-for-freedom-from-virus-restrictions.html

23. SteelFisher GK, Blendon RJ, Ward JRM, Rapoport R, Kahn EB, Kohl KS. Public response to the 2009 influenza A H1N1 pandemic: a polling study in five countries. Lancet Infect Dis. 2012;12(11):845–50.

24. Moon MJ. Fighting Against COVID-19 with Agility, Transparency, and Participation: Wicked Policy Problems and New Governance Challenges. Public Administration Review. 2020.

25. Mouter N, Koster PR, Dekker T. Contrasting the recommendations of Participatory Value Evaluation and Cost-Benefit Analysis in the context of urban mobility investments. Working paper Tinbergen Institute; 2020.

26. Dekker T, Koster P, Mouter N. The economics of participatory value evaluation. SSRN Electron J [Internet]. 2019; Available from: http://dx.doi.org/10.2139/ssrn.3323645.

27. Harari YN. The world after coronavirus. Financial Times [Internet]. 2020 Mar 20 [cited 2020 Nov 3]; Available from: https://www.ft.com/content/19d90308-6858-11ea-a3c9-1fe6fedcca75.

28. Mudde C. “Wartime” coronavirus powers could hurt our democracy – without keeping us safe. The guardian [Internet]. 2020 Mar 24 [cited 2020 Nov 3]; Available from: http://www.theguardian.com/commentisfree/2020/mar/24/wartime-coronavirus-powers-state-of-emergency.

29. Roth K. How authoritarians are exploiting the covid-19 crisis to grab power [Internet]. Nybooks.com. 2020 [cited 2020 Nov 3]. Available from: https://www.nybooks.com/daily/2020/03/31/how-authoritarians-are-exploiting-the-COVID-19-crisis-to-grab-power/.

30. Liu Y, Boin A. Framing a mega-disaster: Political rhetoric and the Wenchuan earthquake. Saf Sci. 2020;125(104621):104621.

31. Capano G, Howlett M, Jarvis DSL, Ramesh M, Goyal N. Mobilizing policy (in)capacity to fight COVID-19: Understanding variations in state responses. Policy Soc. 2020;39(3):285–308.

32. Fiorino, J D. Citizen participation and environmental risk: a survey of institutional mechanisms. Science, Technology and Human Values. 1990;15:226–243.

33. Wright AL, Sonin K, Driscoll J, Wilson J. Poverty and economic dislocation reduce compliance with COVID-19 shelter-in-place protocols. SSRN Electron J [Internet]. 2020; Available from: http://dx.doi.org/10.2139/ssrn.3573637.

34. Webster N. Public discussions on COVID-19 lockdown in Scotland [Internet]. Participo. 2020 [cited 2020 Nov 3]. Available from: https://medium.com/participo/public-discussions-on-COVID-19-lockdown-in-scotland-8f34a586c69c.

35. Esaiasson P, Gilljam M, Persson M. Responsiveness beyond policy satisfaction: Does it matter to citizens? Comp Polit Stud. 2017;50(6):739–65.

36. Gutmann A, Thompson DF. Why deliberative democracy? Princeton University Press; 2004.

37. Mansbridge J. Taking coercion seriously. Constellations. 1997;3(3):407 –16

38. Xafis V. What is Inconvenient for You is Life-saving for Me’: How Health Inequities are playing out during the COVID-19 Pandemic. Asian Bioethics Review. 2020;1

39. Dynes R. Social capital: Dealing with community emergencies. Homeland Security Affairs. 2006;2(2):1–26.

40. Blendon RJ, Koonin LM, Benson JM, Cetron MS, Pollard WE, Mitchell EW, et al. Public response to community mitigation measures for pandemic influenza. Emerg Infect Dis. 2008;14(5):778–86.

41. Allcott H, Boxell L, Conway J, Gentzkow M, Thaler M, Yang D. Polarization and public health: Partisan differences in social distancing during the Coronavirus pandemic. J Public Econ. 2020;(104254):104254

42. Chuang Y-C, Huang Y-L, Tseng K-C, Yen C-H, Yang L-H. Social capital and health-protective behavior intentions in an influenza pandemic. PLoS One. 2015;10(4):e0122970.

43. Bol D, Giani M, Blais A, Loewen PJ. The effect of COVID-19 lockdowns on political support: Some good news for democracy? Eur J Polit Res [Internet]. 2020;(1475-6765.12401). Available from: http://dx.doi.org/10.1111/1475-6765.12401

44. Tufekci Z. How Hong Kong did it. Atlantic monthly (Boston) [Internet]. 2020 May 12 [cited 2020 Nov 3]; Available from: https://www.theatlantic.com/technology/archive/2020/05/how-hong-kong-beating-coronavirus/611524/

45. Hartley K, Jarvis DSL. Policymaking in a low-trust state: legitimacy, state capacity, and responses to COVID-19 in Hong Kong. Policy Soc. 2020;39(3):403–23.

46. Hendriks CM, Lees-Marshment J. Political leaders and public engagement: The hidden world of informal elite–citizen interaction. Polit Stud. 2019;67(3):597–617.

47. Koskimaa V, Rapeli L. Fit to govern? Comparing citizen and policymaker perceptions of deliberative democratic innovations. Policy Polit [Internet]. 2020; Available from: http://dx.doi.org/10.1332/030557320X15870515357288

48. Gilbert M, Dewatripont M, Muraille E, Platteau J-P, Goldman M. Preparing for a responsible lockdown exit strategy. Nat Med. 2020;26(5):643 –4.

49. Rowe G, Frewer LJ. A Typology of Public Engagement Mechanisms. Science, Technology, & Human Values. 2005;30(2):251–290.

50. Goodin RE, Dryzek JS. Deliberative impacts: the macro-political uptake of mini-publics. Politics and society. 2006;34(2):219–244.

51. Pal M. The promise and limits of citizens’ assemblies: deliberation, institutions and the law of democracy. Queen’s LJ. 2012;38:259.

52. Deliberative Democracy Consortium [Internet]. Deliberative-democracy.net. [cited 2020 Nov 3]. Available from: https://deliberative-democracy.net/

53. PPC, University of Nebraska. The Public engagement project on community control measures for pandemic influenza; findings and recommendations from citizen and stakeholder deliberation days. [cited 2020 Nov 3]; Available from: https://stacks.cdc.gov/view/cdc/12171

54. Dryzek JS, Niemeyer S. Discursive representation. American political science review. 2008;120(4):481–492.

55. Goodin RE. Democratic deliberation within. Philos Public Aff. 2000;29(1):81 –109.

56. Frey BS, Stutzer A. Happiness, economy and institutions. Econ J (London). 2000;110(466):918–38.

57. Offe C. Referendum vs. Institutionalized deliberation: What democratic theorists can learn from the 2016 Brexit decision. Daedalus. 2017;146(3):14–27.

58. Lupia A, Matsusaka JG. Direct Democracy: New approaches to old questions. Annu Rev Polit Sci. 2004;7(1):463–82

59. The Big Corona Study (2020), University of Antwerp, University of Hasselt and KU Leuven, www.corona-studie.be

60. Chorus C, Sandorf ED, Mouter N. Diabolical dilemmas of COVID-19: An empirical study into Dutch society’s trade-offs between health impacts and other effects of the lockdown. PLoS One. 2020;15(9):e0238683.

61. Aragones E, Sanchez-Pages S. A theory of participatory democracy based on the real case of Porto Alegre”. European Economic Review. 2009;53:56–72.

62. Cabannes Y. Participatory budgeting: a significant contribution to participatory democracy”. Environment and Urbanization. 2004;16(1):27–46.

63. Sintomer Y, Herzberg C, Rocke A. Participatory Budgeting in Europe: Potentials and Challenges”. International Journal of Urban and Regional Research. 2008;32(1):164 – 178.

64. Hanley N, Mourato S, Wright RE. Choice modelling approaches: a superior alternative for environmental valuation? Journal of Economic Surveys. 2001;15:435 –62.

65. Czajkowski M, Hanley N, LaRiviere J. The effects of experience on preferences: Theory and empirics for environmental public goods. Am J Agric Econ. 2015;97(1):333–51.

66. Bartkowski B, Lienhoop N. Democracy and valuation: A reply to Schläpfer (2016). Ecol Econ. 2017;131:557–60.

67. Dietz T, Stern PC, Dan A. How deliberation affects stated willingness to pay for mitigation of carbon dioxide emissions: An experiment. Land Econ. 2009;85(2):329–47

68. Carson RT, Mitchell RC, Hanemann M, Kopp RJ, Presser S, Ruud PA. Contingent valuation and lost passive use: Damages from the Exxon Valdez oil spill. Environmental and Resource Economics. 2003;25(3):257–286.

69. Carson RT. Contingent valuation: a comprehensive bibliography and history. Edward Elgar Publishing; 2012.

70. Halkos G, Leonti A, Sardianou E. Assessing the preservation of parks and natural protected areas: A review of Contingent Valuation studies. Sustainability. 2020;12(11):4784.

71. Haab TC, McConnell KE. Valuing environmental and natural resources: the econometrics of non-market valuation. Edward Elgar Publishing; 2002.

72. Carson RT, Hanemann WM. Chapter 17 Contingent Valuation. In: Handbook of Environmental Economics. Elsevier; 2005. p. 821–936.

73. Lancaster KJ. A new approach to consumer theory. J Polit Econ. 1966;74(2):132 –57.

74. Hensher DA, Rose JM, Greene WH. Applied Choice Analysis: A Primer. Cambridge: Cambridge University Press; 2005.

75. Train KE. Discrete choice methods with simulation. Cambridge university press; 2009.

76. McFadden D. Conditional logit analysis of qualitative choice behavior. In: Zarembka P, editor. New York: Academic Press; 1974. p. 105–142.

77. Mouter N, van Cranenburgh S, van Wee B. Do individuals have different preferences as consumer and citizen? The trade-off between travel time and safety. Transp Res Part A Policy Pract. 2017;106:333–49

78. Mouter N, Cabral MO, Dekker T, van Cranenburgh S. The value of travel time, noise pollution, recreation and biodiversity: A social choice valuation perspective. Res Transp Econ. 2019;76(100733):100733.

79. Klaassen N. Mag het noorden als eerste uit de lockdown? [Internet]. Parool.nl. 2020 [cited 2020 Nov 3]. Available from: https://www.parool.nl/nieuws/mag-het-noorden-als-eerste-uit-de-lockdown~b369c133/

80. Dutch Health Care Authority, 2020. Heropstart van zorg: eenvoudige rekensommetjes, maar lastige afwegingen. https://www.zorgvisie.nl/heropstart-van-zorg-eenvoudige-rekensommetjes-maar-lastigeafwegingen/

81. The Guardian.com. [cited 2020 Nov 3]. Surge in domestic violence during COVID-19 crisis. Available from: https://www.theguardian.com/society/2020/apr/12/domestic-violence-surges-seven-hundred-per-cent-ukcoronavirus

82. NL Times, 2019. Over 700,000 Dutch domestic violence victims in 5 years. February 6, 2019. https://nltimes.nl/2019/02/06/700000-dutch-domestic-violence-victims-5-years-report

83. NIBUD,2020. Een vijfde van de Nederlanders ervaart inkomensterugval. https://www.nibud.nl/consumenten/nibud-een-vijfde-van-de-nederlanders-ervaart-inkomensterugval/

84. CBP. Scenario’s coronacrisis 26 maart 2020 [Internet]. Cpb.nl. [cited 2020 Nov 3]. Available from: https://www.cpb.nl/scenarios-coronacrisis

85. Rabobank [Internet]. [cited 2020 Nov 3]. Available from: https://economics.rabobank.com/publications/2020/april/dutch-economy-to-contract-this-year-morethan-in-2009/

86. Johnston RJ, Boyle KJ, Adamowicz W (vic), Bennett J, Brouwer R, Cameron TA, et al. Contemporary guidance for stated preference studies. J Assoc Environ Resour Econ. 2017;4(2):319–405

87. Carson RT, Groves T. Incentive and informational properties of preference questions. Environ Resour Econ (Dordr). 2007;37(1):181–210

88. Bhat CR. The multiple discrete-continuous extreme value (MDCEV) model: Role of utility function parameters, identification considerations, and model extensions. Trans Res Part B: Methodol. 2008;42(3):274–303.

89. Reckers-Droog. V. Exel J, Brouwer W. Who should receive treatment? An empirical enquiry into the relationship between societal views and preferences concerning healthcare priority setting. PLoS ONE. 2018;13(6):019876.

90. Wouters S, Exel J, W. Brouwer RB. Priority to end of life treatments? Views of the public in the Netherlands. Value in Health. 2017;20(1):107–117.

91. Aldrich DP, Meyer MA. Social capital and community resilience. Am Behav Sci. 2015;59(2):254–69.

92. Coroiu A, Moran C, Campbell T, Geller AC. Barriers and facilitators of adherence to social distancing recommendations during COVID-19 among a large international sample of adults. PloS one. 2020;15(10).

93. Warren ME, Mansbridge J. Deliberative negotiation. Mansbridge J, Martin CJ, editors. American Political Science Association; 2013.

94. Almaatouq A. Towards stable principles of collective intelligence underan environment-dependent framework [Internet]. PsyArXiv. 2020. Available from: http://dx.doi.org/10.31234/osf.io/ahr7y

95. Almaatouq A, Noriega-Campero A, Alotaibi A, Krafft PM, Moussaid M, Pentland A. Adaptive social networks promote the wisdom of crowds. Proc Natl Acad Sci U S A. 2020;117(21):11379–86.

